# Ensuring anatomical specificity in lesion network mapping: a multicentre study of 2,950 stroke patients

**DOI:** 10.64898/2026.02.16.26346377

**Authors:** Marvin Petersen, Kaustubh R. Patil, Simon B. Eickhoff, Geert Jan Biessels, the Meta VCI Map Consortium

## Abstract

As many neurobehavioural functions rely on distributed brain networks, anatomically diverse brain lesions can cause highly similar neurobehavioural deficits. Lesion network mapping (LNM) builds on this principle to identify symptom-specific brain networks by mapping focal lesions onto a normative functional connectome. Recent work, however, questioned the anatomical specificity of LNM, showing that commonly used LNM procedures converge on nonspecific properties of the normative connectome rather than symptom-specific networks. Here, we investigated whether anatomically specific LNM can be achieved through appropriate study design and statistical inference. Using a multicentre dataset of 2,950 stroke patients across 12 cohorts, we compared lesion-derived connectivity maps between patients with and without impairment across six cognitive domains. We evaluated group differences using both permutation of symptom labels and parametric two-sample inference. Additionally, we examined a one-sample approach restricted to cognitively impaired patients, i.e., without an unimpaired comparison group, similar to what was used in the previous critique. Both two-sample approaches yielded distinct, biologically plausible lesion network maps with evident domain-specific features, and limited correspondence to nonspecific connectome properties. Their cross-domain similarity followed both the co-occurrence of deficits across domains and the similarity of connectome-independent voxel-based lesion-symptom maps derived from the same lesions, consistent with overlap and divergence between maps being driven by the covariance structure of the cognitive data. Simulation analyses across 10,000 null studies confirmed valid type I error control of the permutation framework. In contrast, the one-sample analyses, which aggregated lesion connectivity maps across affected patients without a control comparison, reproduced the previously described convergence toward nonspecific connectome properties and showed marked similarity across cognitive domains. These findings demonstrate that connectome-driven convergence is not an inherent limitation of LNM, but depends critically on study design: case-control comparisons with appropriate statistical inference ensure anatomical specificity and provide a robust framework for identifying brain networks associated with distinct neurobehavioural deficits.

## Introduction

That anatomically distinct lesions can cause highly similar neurobehavioural symptoms has been a longstanding enigma in neuroscience. One explanation is that distinct lesions may disrupt brain areas belonging to a common brain network: a lesion anywhere in a network fundamental to a given function will impair that function, regardless of the exact lesion site. Lesion network mapping (LNM) operationalizes this principle, leveraging this many-to-one relationship to offer a network perspective on the brain’s functional anatomy.^1^ By projecting lesion locations onto a normative functional connectome, LNM generates lesion network maps that help to identify the distributed neural circuits underlying clinical deficits.^2^ This premise has contributed to its rapid adoption across neurology and psychiatry.

However, recent work has raised concerns about the anatomical specificity of LNM.^3^ Across multiple analyses, lesion network maps derived from clinically distinct symptoms were observed to exhibit substantial similarity, suggesting that commonly used LNM procedures may preferentially recover general properties of the underlying normative connectome rather than symptom-specific brain networks. The proposed explanation was that lesion-derived connectivity maps are inherently shaped by the fixed connectivity architecture onto which all lesions are projected, introducing a connectome-driven bias whereby features such as regional hubness or degree dominate statistical maps. These concerns have been raised across different implementations of LNM, including approaches that aggregate lesion-derived connectivity maps across affected patients (often referred to as *sensitivity analyses*) as well as symptom-anchored analyses relating patient-level connectivity profiles to clinical measures or comparing them between cases and controls (*specificity analyses*).^3^ While these observations have prompted important questions regarding the specificity and interpretability of LNM, the extent to which they generalize across different analytical frameworks remains an active topic of debate.

These considerations prompted us to revisit the methodological approach used in our own earlier work applying LNM to white matter hyperintensity data,^4^ which relied on permutation of symptom labels. In principle, label permutation preserves the structured dependence induced by the normative connectome while breaking the relationship between lesion-derived connectivity maps and the clinical phenotype, such that connectome-driven convergence is incorporated into the null distribution. Permutation-based inference of this kind is well established in neuroimaging and has been implemented in several lesion network mapping studies.^5–8^ However, whether this inferential framework – or a symptom-anchored case-control design more generally – mitigates the described convergence has not been evaluated systematically.

Hence, the central unresolved question we address here is whether appropriate study design and statistical inference can distinguish nonspecific connectome properties from symptom-specific network effects.^9^ We therefore tested whether anatomically specific lesion network maps can be achieved within a symptom-anchored case-control framework. To do so, we used a multicentre dataset of 2,950 patients with acute ischemic stroke from the Meta VCI Map Consortium. Acute ischemic stroke provides a prototypical model of focal lesions which underpin much of the lesion network mapping literature. We compared three analytical strategies, which have been previously used in this context: 1) label permutation^5^ and 2) parametric two-sample inference^10^, both contrasting cognitively impaired and unimpaired patients in a case-control design, and 3) one-sample aggregation of lesion-derived connectivity maps only taking impaired patients into account.^3^ We further evaluated the validity of the label permutation framework through simulation analyses and related cross-domain similarity of the resulting lesion network maps to deficit co-occurrence and to cross-domain similarity of connectome-independent voxel-based lesion-symptom maps. By systematically comparing these approaches, we show that symptom-anchored case-control analyses yield anatomically specific and biologically plausible lesion network maps with both label permutation-based and parametric inference, whereas one-sample analyses reproduce the connectome-driven convergence described previously.^3^

## Materials and methods

We analysed data from 2,950 stroke patients across 12 post-stroke cognitive impairment cohorts of the Meta VCI Map Consortium, all with acute ischemic stroke lesion segmentations and harmonized cognitive assessment.^11^

Neuropsychological assessment was performed within one year of the index stroke. Cognitive status was classified as impaired or unimpaired for six domains – attention/executive function, processing speed, language, verbal memory, visuospatial functions, and visuospatial memory. Each domain aggregates performance across several standardized neuropsychological tests, grouped according to the harmonization procedure established within the Meta VCI Map Consortium, with impairment defined as performance below the fifth percentile of local normative data within each contributing cohort. The domains are therefore multi-test composites rather than single isolated cognitive processes; the full per-test operationalization is given in the source description of the dataset.^11^

Acute infarcts were segmented within each cohort and normalized to MNI152NLin6Asym space. Lesion-derived functional connectivity maps, estimating the brain circuitry connected to each patient’s lesion, were computed in Lead-DBS using the preprocessed GSP1000 normative functional connectome.^12,13^ This connectome comprises resting-state fMRI data of 1,000 healthy young adults, preprocessed with a modified version of the Computational Brain Imaging Group pipeline (https://github.com/bchcohenlab/BIDS_to_CBIG_fMRI_Preproc2016). Each patient’s binary lesion mask served as a seed region. For each GSP1000 participant, the mean blood-oxygen-level-dependent (BOLD) time course across all lesion voxels was correlated with the time course of every brain voxel, and the correlation coefficients were Fisher z-transformed. The z-values were then summarized voxelwise across the 1,000 participants as a T-statistic (mean z-value divided by its standard error), reflecting how consistently each voxel is functionally connected to the lesion across the normative sample. The resulting unthresholded maps served as input for all group-level analyses. Following the established lesion network mapping protocol, lesion seeds and target voxels were not restricted to grey matter.

Statistical analyses were conducted with Permutation Analysis of Linear Models (PALM) using model specifications matched as closely as possible across inference approaches.^14^ For each cognitive domain, inference was performed using three approaches: (i) symptom-label permutation testing contrasting cognitively impaired and unimpaired patients, (ii) a parametric two-sample t-test contrasting cognitively impaired and unimpaired patients, and (iii) a one-sample t-test restricted to cognitively impaired patients. Each voxelwise statistical map was corrected for multiple comparisons using the false discovery rate (FDR). Label permutation-based analyses employed 1,000 permutations with threshold-free cluster enhancement (TFCE) and p-values were calculated as the proportion of permuted test statistics equalling or exceeding the observed statistic (two-sided).^14^ One-sample t-tests were likewise performed with TFCE, using sign-flipping (1,000 permutations) to generate the null distribution. Because patients were pooled across the 12 cohorts, cohort was modelled as a fixed effect, and label permutation-based inference was performed within cohort using exchangeability blocks. Primary label permutation, parametric group analyses and one-sample t-tests were performed without further covariates beyond cohort. For all three inference approaches, models additionally adjusting for lesion volume and NIHSS are reported in the supplementary materials. Missing NIHSS values (n = 296) were median-imputed.

To quantify pairwise similarity across cognitive domains, we computed spatial correlations of unthresholded T-statistic maps and Dice overlap of thresholded significance maps. Pairwise similarity with the degree map of the GSP1000 connectome was also quantified. Degree map alignment of one-sample and case-control maps was compared by a paired t-test on Fisher-z-transformed spatial correlations across the six domains.

Prior lesion network mapping sensitivity analyses and the recent critique aggregated lesion-derived connectivity maps across affected patients without a control comparison, for example by voxelwise averaging or by overlap of thresholded maps.^3^ The one-sample t-test used here formalizes this aggregation as an inference step comparable to the two case-control approaches. To assess whether the two differ, we compared one-sample T-maps and mean-aggregated maps via spatial correlation within each domain.

For the label permutation-based lesion network maps, domain-specific effects were anatomically localized by identifying the five strongest positive local maxima (peak voxels) in each T-statistic map, constrained to be at least 20 mm apart. Peaks were assigned to atlas parcels (Schaefer1000 cortex, Tian54 subcortex, and Buckner cerebellum), with peaks located in white matter assigned to the nearest grey matter parcel.^15–17^ To quantify peak specificity, the T-statistic and FDR significance at each of these 30 peak voxels (five per domain across six domains) were extracted from all six lesion network maps, and the own-domain T-statistic was compared with the mean absolute T-statistic of the other five domains.

To assess type I error control of the label permutation framework under the null hypothesis, we conducted simulation analyses across 10,000 null studies using both synthetic lesions and real lesion masks from the Meta VCI Map cohort. Under a valid null, the proportion of significant ROIs (p < 0.05) should approximate the nominal false positive rate (≈5%). We quantified the empirical false positive rate and characterized its spatial distribution and associations with nonspecific connectome properties (degree and PC1). Details are presented in the supplementary materials.

To examine the spatial correspondence between label permutation-based lesion network maps and conventional lesion-symptom associations, we further compared the resulting networks with voxel-based lesion-symptom mapping (VLSM) performed on the same dataset taking binary lesion masks as input. VLSM was performed within the same statistical framework as label permutation-based LNM: a voxelwise general linear model in PALM contrasting impaired and unimpaired patients for each cognitive domain, with symptom-label permutation (1,000 permutations, two-sided), TFCE, and FDR correction. VLSM analyses were covariate-matched to the LNM analyses: the primary model included cohort as the only covariate, and a fully adjusted model additionally included lesion volume and NIHSS. VLSM thus differed from LNM only in the projection of lesions onto the normative connectome, the step proposed to render lesion network maps similar across symptoms.^3^ If this projection drove map similarity, lesion network maps should converge across domains while the corresponding VLSM maps remain distinct; conversely, if label permutation merely removed the nonspecific connectome signal shared by all patients, lesion network maps should reduce to the lesion-symptom anatomy captured by VLSM. To assess whether the correspondence between both methods was domain-specific, voxelwise correlations were computed between all lesion network and VLSM T-maps, and same-domain pairs were compared with different-domain pairs. Throughout, LNM refers to label permutation-based lesion network mapping unless stated otherwise.

Pairwise co-occurrence of the binary impairment labels was quantified as phi coefficient and Jaccard index. Associations between cross-domain similarity matrices (lesion network maps, VLSM maps, and impairment co-occurrence) were assessed by Spearman correlation with Mantel tests (exact and two-sided). To test whether cross-domain similarity of lesion network maps reflects co-occurring deficits, the label permutation analysis for attention/executive function, language, and verbal memory was repeated with the binary impairment labels of the other two domains as additional covariates, restricted to patients with complete data on all three domains.

Each contributing cohort obtained local ethics approval and written informed consent, as previously described.^11,18^

## Results

Sample characteristics are summarized in ***Table 1***. Lesion coverage in the dataset was high, with 86.9% of brain voxels affected in at least five patients. For a lesion distribution heatmap see ***Supplementary figure S1***. Average pairwise co-occurrence of cognitive domain impairment was low (mean phi = 0.30 ± 0.10, mean Jaccard index = 0.26 ± 0.08; ***Supplementary figure S2***), showing that patients had diverse deficits.

**Table 1.** Sample characteristics.

| Age, years, mean ± SD | 66.8 ± 11.6 |
|---|---|
| Female patients, n (%) | 1,157 (39.2) |
| **Education** |  |
| Lower than secondary school, n (%) | 1,703 (57.7) |
| Secondary school, n (%) | 608 (20.6) |
| Technical school or college, n (%) | 234 (7.9) |
| University or higher, n (%) | 405 (13.7) |
| **Ethnicity** |  |
| Korean, n (%) | 1,394 (47.3) |
| European, n (%) | 1,069 (36.2) |
| Chinese, n (%) | 462 (15.7) |
| Malay, n (%) | 16 (0.5) |
| Indian, n (%) | 6 (0.2) |
| Unknown, n (%) | 3 (0.1) |
| **Clinical characteristics** |  |
| Baseline NIHSS, median [IQR] | 3 [1–5] |
| Cognitive assessment timing, days after index stroke event, median [IQR] | 105 [74–170] |
| Clinical history of stroke, n (%) | 327 (11.1) |
| **Post-stroke cognitive impairment** |  |
| Any-domain, n<sub>impaired</sub> / n<sub>available</sub> (%) | 1,181 / 2,340 (50.5) |
| Attention / executive function, n<sub>impaired</sub> / n<sub>available</sub> (%) | 375 / 2,192 (17.1) |
| Processing speed, n<sub>impaired</sub> / n<sub>available</sub> (%) | 360 / 2,088 (17.2) |
| Language, n<sub>impaired</sub> / n<sub>available</sub> (%) | 372 / 2,301 (16.2) |
| Verbal memory, n<sub>impaired</sub> / n<sub>available</sub> (%) | 605 / 2,283 (26.5) |
| Visuospatial functions, n<sub>impaired</sub> / n<sub>available</sub> (%) | 509 / 1,872 (27.2) |
| Visuospatial memory, n<sub>impaired</sub> / n<sub>available</sub> (%) | 98 / 806 (12.2) |
| **Brain imaging** |  |
| Normalized acute infarct volume, mL, median [IQR] | 2.7 [1.0–14.1] |
| Imaging timing, days after event, median [IQR] | 4 [1–9] |

NIHSS = National Institutes of Health Stroke Scale; IQR = interquartile range; SD = standard deviation.

Label permutation yielded statistically significant lesion network maps for five of the six domains (***Figure 1***; for unthresholded maps see ***Supplementary figures S3 and S4***): attention/executive function, processing speed, language, verbal memory, and visuospatial functions; visuospatial memory showed no significant group differences under the permutation null. Parametric two-sample t-tests produced per-domain lesion network maps that were highly comparable to those from the label permutation analysis (***Supplementary figures S5–S7***, mean Dice = 0.85 ± 0.05). Additionally adjusting for lesion volume and NIHSS also resulted in highly comparable lesion network maps, although visuospatial functions no longer reached significance (***Supplementary figure S8***). For parsimony, the following paragraphs report as the primary analysis the label permutation model adjusted for cohort only, i.e., without lesion volume and NIHSS. Unless stated otherwise, unqualified references to any of the inference approaches hereafter denote the respective model adjusted for cohort only.

**Figure 1.**
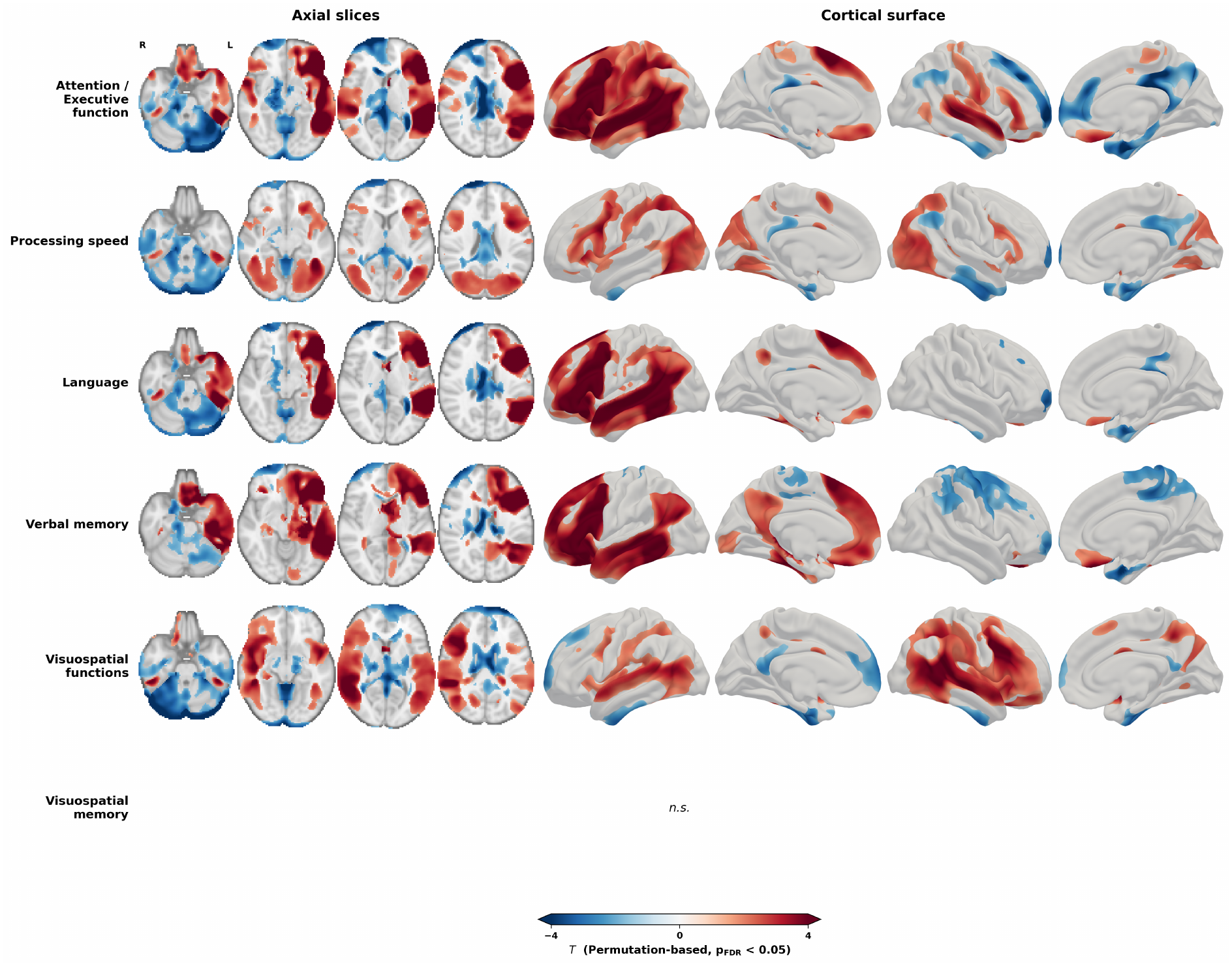
Label permutation-based lesion network maps on axial slices. Lesion network maps across the six cognitive domains, thresholded at p_FDR_ < 0.05 and shown on four axial slices (z = −26, −10, 6, 22 mm; radiological orientation) as well as cortical surface renderings (Conte69 midthickness). Colours denote the direction of the group difference in lesion-derived functional connectivity: red, voxels more strongly connected to the lesions of impaired than unimpaired patients (impaired > unimpaired); blue, voxels more strongly connected to the lesions of unimpaired patients (impaired < unimpaired). Red thus marks the network whose disruption is associated with impairment in each domain. No voxels survived for visuospatial memory (n.s.). The colour scale is shared across panels. yabplot v0.5.1 was used for volume- to-surface projection and visualization.^27^

Across the five domains with significant maps, the label permutation-based maps overlapped only weakly with the connectome’s degree map (mean Dice = 0.20 ± 0.07, mean r = 0.28 ± 0.24) and were only moderately similar to one another (mean Dice = 0.50 ± 0.11, mean r = 0.55 ± 0.28) (***Figure 2***), showing that the identified networks did not converge onto a shared, domain-general pattern driven by nonspecific connectome structure. Anatomical specificity was most apparent for language, where the map highlighted left-lateralized perisylvian regions, including inferior frontal and temporal cortex, as expected a priori. Notably, the three left-dominant maps were differentiated beyond their shared left frontotemporoparietal core: the attention/executive function map additionally included right-hemispheric regions, whereas the verbal memory map featured midline default mode network structures and more prominent hippocampal and parahippocampal involvement.

**Figure 2.**
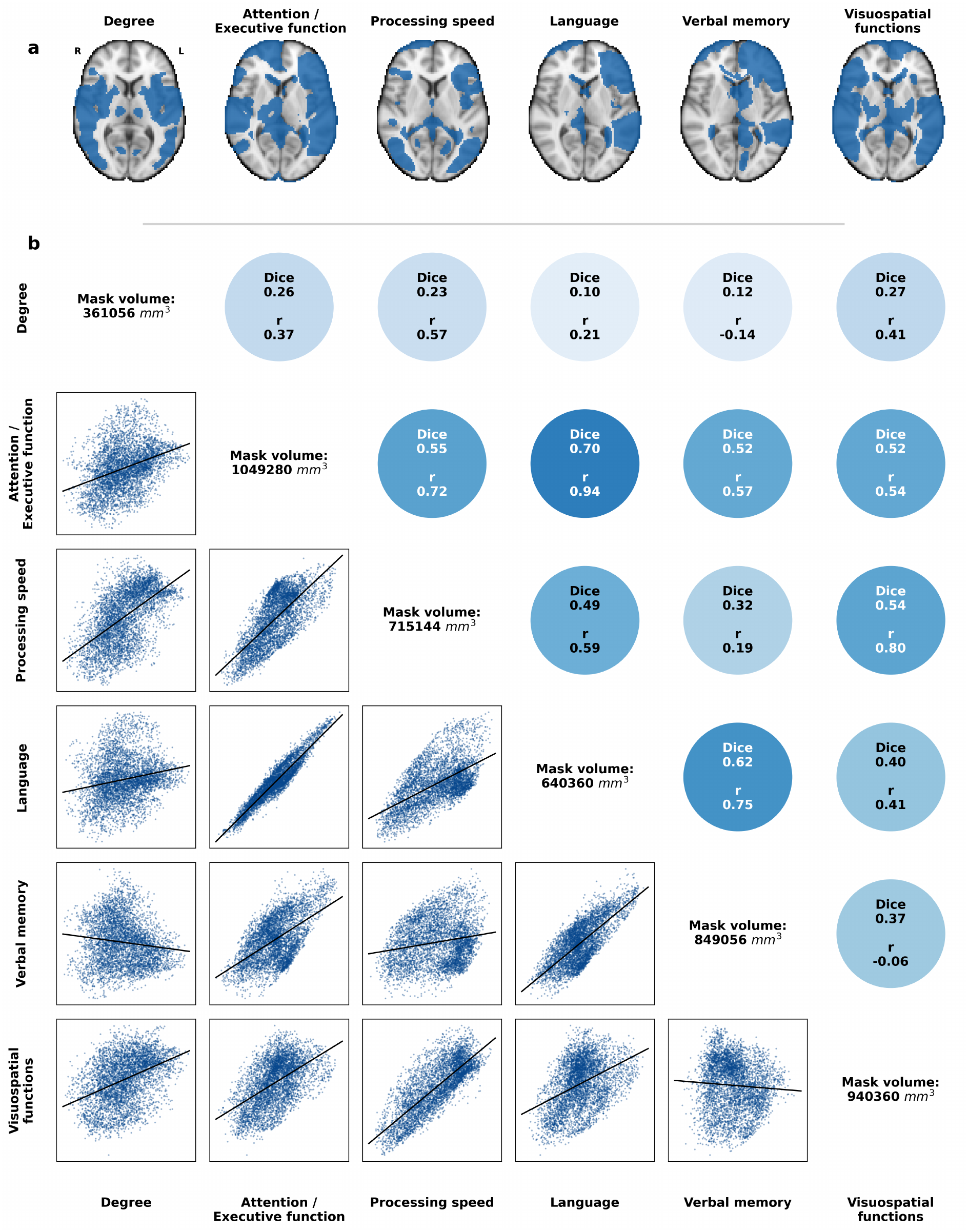
Spatial correspondence of label permutation-based lesion network maps. (a) Symptom-linked lesion network maps derived using label permutation, thresholded at p_FDR_ < 0.05. Visuospatial memory is not shown, as no voxels survived thresholding. (b) Pairwise comparisons of label permutation-based lesion network maps. Upper triangle: similarity quantified as Dice overlap of significance-thresholded maps. The first row/column compares each map with the normative connectome degree map (degree defined as the row-sum of the normative FC matrix; binary degree mask thresholded at the 95th percentile). Lower triangle: pairwise spatial correlation illustrated by regression plot of voxels from the unthresholded T-statistic maps. Diagonal: volumes of thresholded masks of degree and significance maps (mm³).

Peaks of the T-statistics underlying both label permutation and parametric inference differed across cognitive domains, underscoring map specificity (***Figure 3a***). The attention/executive function, processing speed, language, and verbal memory maps were left-lateralized, with all top peaks located in the left hemisphere, whereas visuospatial functions peaked exclusively in the right hemisphere; visuospatial memory peaks were bilaterally distributed. In every domain, the strongest peak fell in white matter proximal to a cortical or subcortical parcel. Attention/executive function peaked in white matter proximal to the left posterior parietal cortex (dorsal attention network, T = +8.3), the same area that led the language map (T = +7.5). Processing speed showed its strongest significant peak in white matter proximal to the left lateral PFC (control network, T = +4.3), verbal memory in white matter proximal to the left caudate (T = +7.7), and visuospatial functions in white matter proximal to the right lateral PFC (control network, T = +5.4). Quantitatively, peaks reached significance in other domains as well (***Figure 3b***). However, the own-domain T-statistic exceeded the mean T-statistic of the other domains at 29 of 30 peaks (mean T = 5.6 vs. mean |T| = 2.7; ***Figure 3c***).

**Figure 3.**
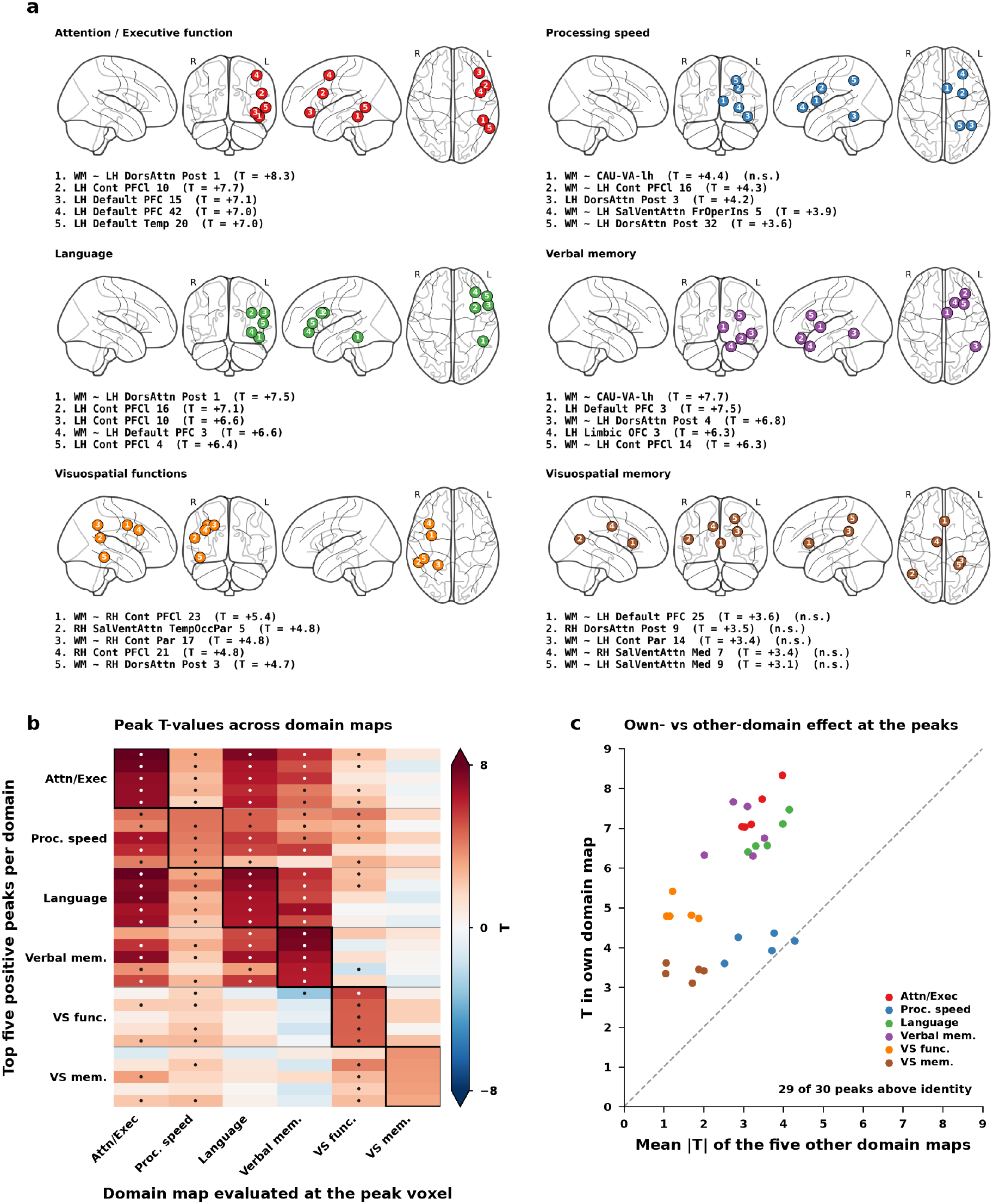
Top five positive T-statistic peaks of lesion network maps and their specificity across cognitive domains. (a) For each domain, the five strongest positive local maxima (≥20 mm apart as minimum separation constraint) of the unthresholded impaired-versus-unimpaired map are shown on a glass brain (left to right: sagittal right hemisphere, coronal, sagittal left hemisphere, axial), numbered by rank, and listed with their nearest anatomical parcel and T-statistic (R, right; L, left; n.s., peak not significant at p_FDR_ < 0.05 in its own domain map). Anatomical labels follow the Schaefer1000×7 cortical, Tian54 subcortical, and Buckner cerebellar parcellations; peaks falling in the white matter are labelled by their nearest parcel. (b) Signed T-statistics of the 30 peak voxels (rows, five per domain) in each of the six label permutation-based lesion network maps, evaluated at the identical MNI voxel (columns). Black outlines mark each peak voxel set’s own domain; dots mark voxels significant at p_FDR_ < 0.05. (c) Each peak’s T-statistic in its own domain map against the mean absolute T-statistic of the five other domain maps at the same voxel, coloured by domain. The dashed line represents identity. The absolute value is used for the other domains so that effects of opposite sign count in full towards the comparison. Attn/Exec, attention/executive function; Proc. speed, processing speed; Verbal mem., verbal memory; VS func., visuospatial functions; VS mem., visuospatial memory.

In contrast to label permutation-based (case-control) lesion network maps, in the same dataset the one-sample t-tests of cases generated maps that corresponded to a higher extent with the normative degree map (mean r = 0.69 ± 0.02; mean Dice = 0.31 ± 0.01) and showed high similarity across the different cognitive domains (mean r = 0.98 ± 0.01; mean Dice = 0.94 ± 0.03; ***Figure 4a***, ***Supplementary figures S9 and S10***). For example, the one-sample t-test language map lacked the expected left lateralization and aligned with the normative degree map (r = 0.70). The difference in degree map alignment between one-sample and case-control maps was statistically significant (paired t-test on Fisher-z-transformed correlations across the six domains: mean r = 0.69 vs. 0.29, t(5) = 4.92, p = 0.004; ***Figure 4b***). Own-domain correspondence between one-sample and case-control maps was low to modest (r = −0.11 to 0.49; ***Figure 4c*** and ***Supplementary figure S11***). One-sample t-tests additionally adjusted for lesion volume and NIHSS, and voxelwise averaging of the connectivity maps across impaired patients, yielded highly comparable maps (voxelwise r ≥ 0.99 and r = 0.91–0.93 against the one-sample T-maps only adjusted for cohort, respectively; ***Supplementary figures S12–S14***). Overlap between label permutation-based and one-sample t-test significance maps was moderate (mean Dice = 0.56 ± 0.07, ***Supplementary figure S15***).

**Figure 4.**
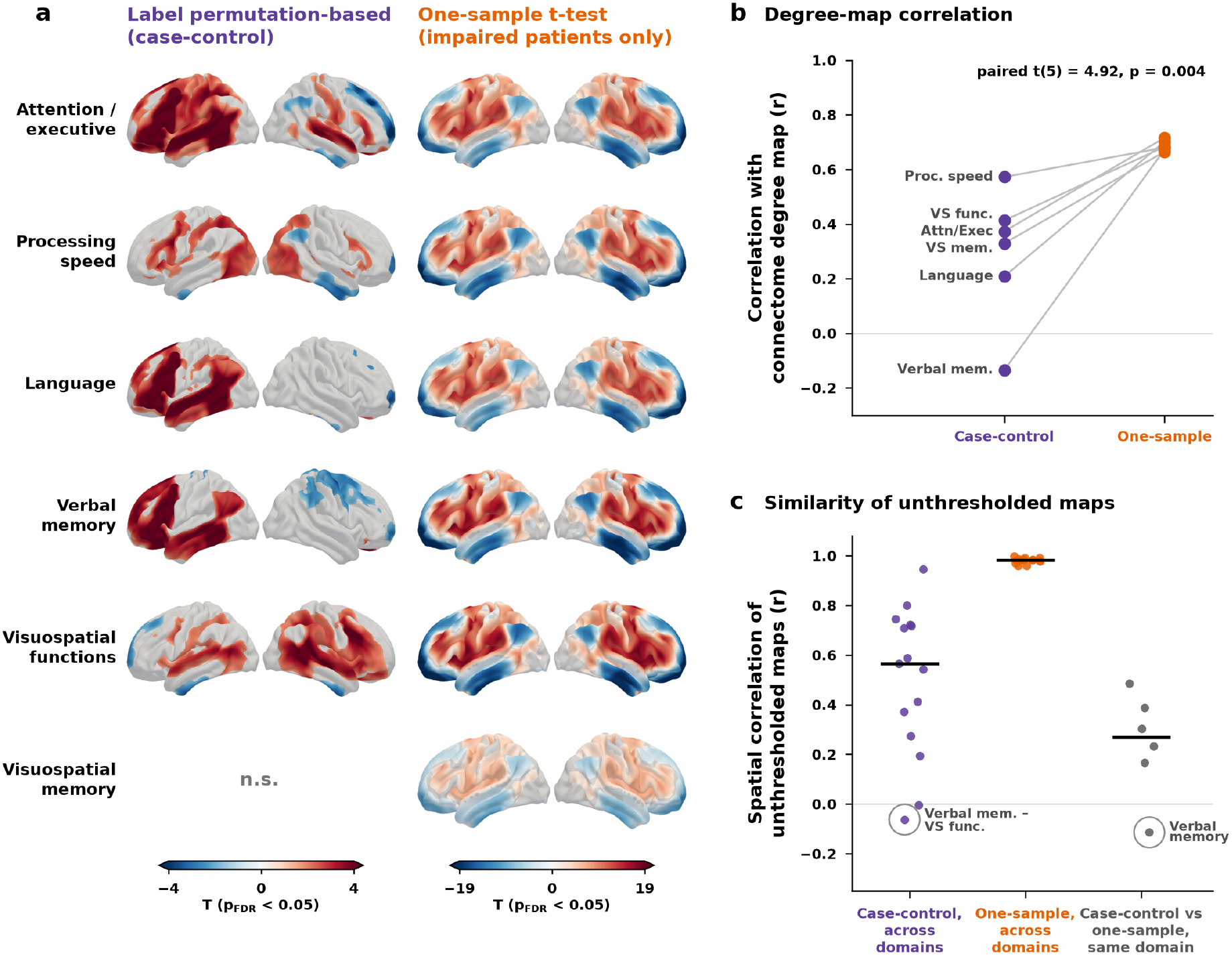
Correspondence between one-sample and case-control lesion network mapping. (a) Label permutation-based lesion network maps (case-control design) and one-sample t-test maps of cognitively impaired patients for the six cognitive domains, thresholded at p_FDR_ < 0.05 and shown on lateral views of the left and right hemisphere; colour scales differ between designs. No voxels survived thresholding in the label permutation-based map for visuospatial memory (n.s.). (b) Spatial correlation of each domain map with the normative connectome degree map for the case-control and one-sample maps. Lines connect the same cognitive domain. Paired t-test on Fisher-z-transformed correlations across the six domains. (c) Spatial correlation of the unthresholded T-maps between case-control maps of different domains, between one-sample maps of different domains, and between the case-control and one-sample maps of the same domain. Horizontal lines mark medians. Circles around dots mark examples. Attn/Exec, attention/executive function; Proc. speed, processing speed; Verbal mem., verbal memory; VS func., visuospatial functions; VS mem., visuospatial memory.

Simulation analyses confirmed valid type I error control under permutation testing across 10,000 null studies. Combining lesion masks with randomly generated symptom scores, the observed mean false positive rate was close to the nominal 5% (5.0% for synthetic and 5.1% for real lesions; ***Supplementary figure S16***). Following the simplified, region-level formulation of the previous critique,^3^ adopted here for computational efficiency, raw symptom-weighted lesion network maps aligned with connectome degree and PC1, reproducing the convergence described previously. Applying label permutation markedly reduced this alignment.

Label permutation-based lesion network maps overlapped only partially with the covariate-matched voxel-based lesion-symptom maps of the same cognitive domain (***Figure 5a***; ***Supplementary figures S17–S20***). The mean Dice coefficient was 0.46 ± 0.04 in the models adjusted for cohort only and 0.48 ± 0.07 in the models additionally adjusted for lesion volume and NIHSS. The unthresholded T-maps of the same domain showed a comparable degree of correspondence, with a mean voxelwise correlation of r = 0.52 in the cohort-only and r = 0.49 in the fully adjusted models (***Figure 5b–c***; ***Supplementary figure S21***). Hence, the two approaches captured partially distinct spatial information. Nonetheless, this correspondence was domain-specific: each lesion network map correlated most strongly with the voxel-based lesion-symptom map of its own domain, whereas correlations between maps of different domains were considerably lower (mean r = 0.27 in the cohort-only and 0.20 in the fully adjusted models; ***Figure 5b–c***). The voxel-based lesion-symptom maps varied in their similarity across domains (***Supplementary figure S22***). Cross-domain similarity of lesion network maps closely followed that of the voxel-based lesion-symptom maps (Spearman ρ = 0.76, Mantel p = 0.001; ***Figure 5d***). For both approaches, map similarity tracked the co-occurrence of the corresponding impairments across patients (lesion network maps: ρ = 0.71, p = 0.001; voxel-based lesion-symptom maps: ρ = 0.74, p = 0.004; ***Figure 5e***).

**Figure 5.**
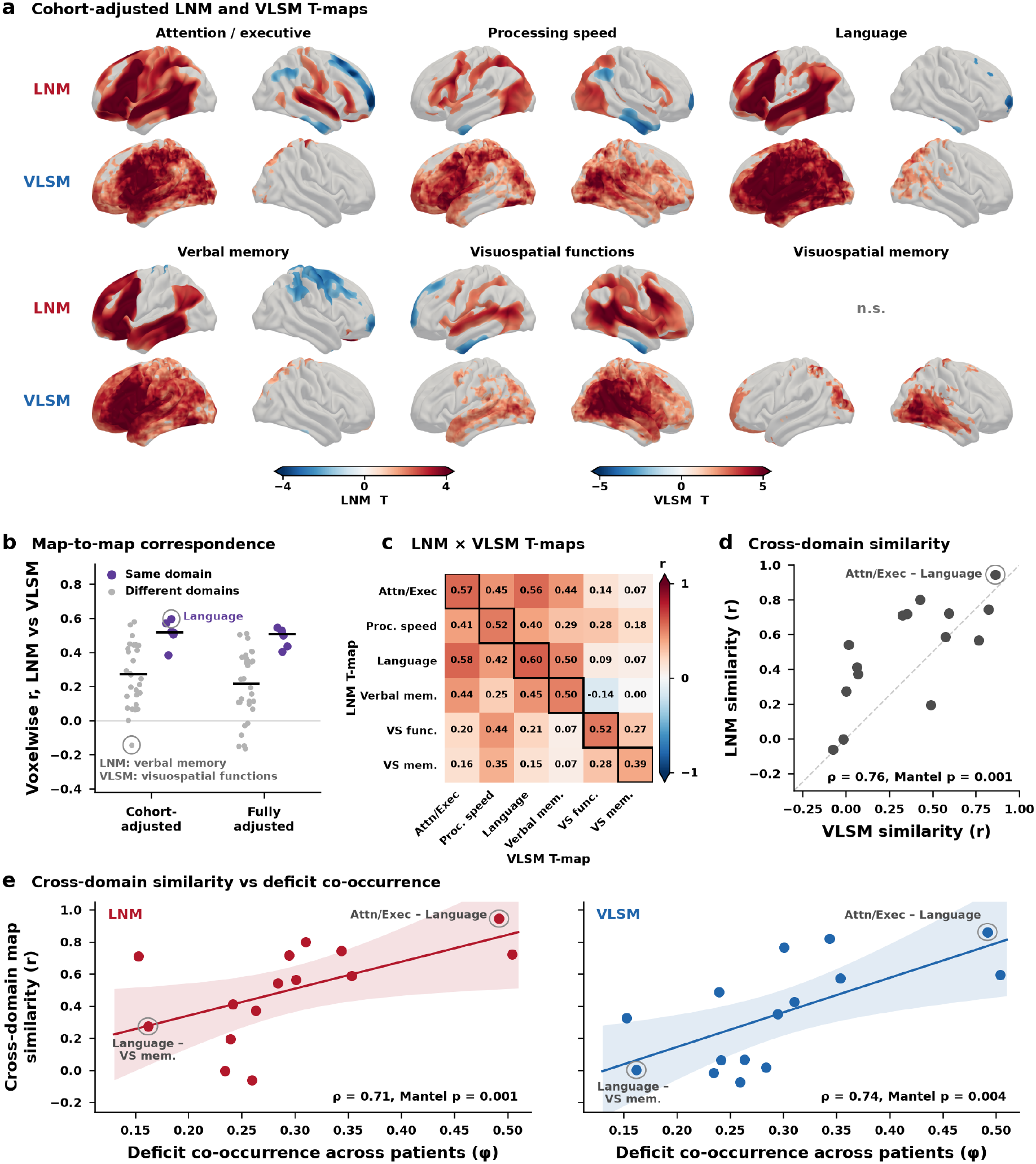
Correspondence between lesion network mapping and voxel-based lesion-symptom mapping. (a) Label permutation-based lesion network maps (LNM) and voxel-based lesion-symptom maps (VLSM) for the six cognitive domains (cohort as the only covariate in both analyses), thresholded at p_FDR_ < 0.05 and shown on lateral views of the left and right hemisphere. LNM and VLSM maps are shown on separate colour scales. (b) Voxelwise correlation between the LNM and VLSM T-maps of the same domain (purple) and of different domains (grey), for the cohort-only and fully adjusted models. Horizontal lines mark medians. Circles highlight one same-domain and one different-domain example. (c) Voxelwise correlations between the LNM T-maps (rows) and the VLSM T-maps (columns), cohort-only models. Black outlines mark same-domain pairs. (d) Cross-domain similarity (Pearson r of the unthresholded T-maps) of LNM plotted against that of VLSM for the 15 domain pairs. The dashed line indicates identity. (e) Cross-domain map similarity as a function of the co-occurrence of the two impairments across patients (phi coefficient), shown separately for LNM (left) and VLSM (right); solid lines and shaded areas show least-squares fits with 95% confidence bands. In d and e, correlations are Spearman ρ with exact two-sided Mantel tests. Attn/Exec, attention/executive function; Proc. speed, processing speed; Verbal mem., verbal memory; VS func., visuospatial functions; VS mem., visuospatial memory.

Subsequently we assessed effects of directly adjusting for co-occurrence of cognitive deficits. This analysis was restricted to attention/executive function, language, and verbal memory, as these domains showed higher spatial overlap. When additionally adjusted for the impairment labels of the other two domains, lesion network maps for attention/executive function, language, and verbal memory retained their own topography (voxelwise correlation with the original unthresholded lesion network map: r = 0.93 attention/executive, r = 0.81 language, r = 0.87 verbal memory), while the similarity among the three maps decreased (attention/executive ∼ language: r = 0.94 to 0.38, attention/executive ∼ verbal memory: r = 0.57 to −0.26; language ∼ verbal memory: r = 0.75 to 0.41; ***Supplementary figure S23***). The adjusted language map no longer reached significance.

## Discussion

In this multicentre study of 2,950 stroke patients, we demonstrated that lesion network mapping produces anatomically specific and biologically plausible network maps when performed within a symptom-anchored case-control framework. Lesion network maps derived from comparisons between cognitively impaired and unimpaired patients showed anatomically plausible patterns such as the expected left-lateralized language network, only moderate similarity across cognitive domains, and little correspondence with nonspecific connectome properties. Importantly, this specificity was achieved using non-parametric label permutation as well as with parametric two-sample inference. In contrast, one-sample analyses that aggregated lesion connectivity maps from impaired patients alone reproduced the strong convergence across symptoms and the alignment with connectome hubs described previously.^3^

Our findings help reconcile recent methodological concerns regarding lesion network mapping. It has recently been suggested that the reported convergence of lesion network maps primarily reflects analyses that summarize lesion connectivity across affected patients (often termed sensitivity analyses), rather than analyses that explicitly compare patients with and without a symptom (specificity analyses).^19^ Our results support this distinction and suggest that the recently described convergence is not an inherent property of lesion network mapping itself, but depends critically on study design and statistical inference. Lesion-derived connectivity maps share variance because all lesions are projected onto the same normative connectome. However, this shared variance is present in both patients with and without a given symptom. Consequently, comparing lesion connectivity between clinically defined groups allows this shared background signal to be distinguished from symptom-related effects. By contrast, one-sample analyses that simply aggregate lesion connectivity maps across affected patients lack such a reference and therefore converge toward highly connected regions.

Both permutation of symptom labels and parametric two-sample testing produced specific lesion network maps in the present dataset. The concordance between these approaches suggests that the primary determinant of anatomical specificity is the symptom-anchored case-control design itself rather than the inference approach. Although the resulting maps were highly similar, the two approaches differ in their underlying statistical assumptions. Parametric inference assumes independent observations, whereas lesion-derived connectivity maps are inherently correlated through the shared normative connectome. Whether this assumption is met by lesion-derived connectivity maps cannot be established from the present data alone. Permutation-based inference, which has been widely adopted in lesion network mapping studies, provides an alternative framework that does not require this independence assumption.^14^ The accompanying simulation analyses further support the validity of label permutation by demonstrating nominal type I error control across 10,000 null studies while reducing alignment with dominant connectome properties.

These findings also have important implications for interpreting the existing lesion network mapping literature. Symptom-anchored case-control analyses – whether analysed with label permutation or parametric inference – are not a new methodological proposal but have already been implemented in many previous LNM studies.^5–8,10,20–23^ Our results therefore provide empirical support for this body of work, suggesting that these studies are unlikely to be explained solely by nonspecific connectome properties. More broadly, our findings complement recent methodological efforts to evaluate lesion network mapping across different study designs, including the development of dedicated null models for case-only analyses.^9^ Together, these developments indicate that connectome-driven convergence can be addressed using principled study design and statistical inference rather than representing an inherent limitation of the technique.^9,19,24^

Our additional analyses further support the biological relevance of the identified networks. We applied voxel-based lesion-symptom mapping with the identical statistical model, impairment labels and inference to the same lesions, such that it differed from lesion network mapping only in the projection onto the normative connectome, the step proposed to render lesion network maps similar across symptoms.^3^ If this projection were the primary determinant of cross-domain similarity, the similarity between lesion network maps of two domains should be high regardless of the similarity between the corresponding lesion-symptom maps. Instead, cross-domain similarity of lesion network maps varied and closely followed that of the connectome-independent lesion-symptom maps: domain pairs with similar lesion-symptom maps also had similar lesion network maps. At the same time, each lesion network map diverged in its topography from the lesion-symptom map of the same domain while aligning only weakly with connectome degree, indicating a symptom-specific network topography that reduces neither to lesion location nor to the hub architecture of the connectome. Previous work suggests that such network-level information improves individual-level symptom prediction over lesion location.^4,25,26^

The shared similarity structure of both map types points to a common source outside the connectome: the co-occurrence of cognitive deficits. Similarity between maps of different domains tracked how often the corresponding impairments co-occurred in the same patients, to a comparable degree for lesion network and lesion-symptom maps. When the three spatially most overlapping domains were additionally adjusted for each other’s impairment labels, their lesion network maps retained their own topography, whereas their mutual similarity decreased markedly. Part of the overlap between lesion network maps of different symptoms thus reflects deficits that co-occur in the same patients rather than a lack of anatomical specificity. Once this shared behavioural variance was removed, the verbal memory network, centred on hippocampal, parahippocampal and midline default mode regions, resembled the human memory circuit derived from lesions causing amnesia.^20^ These findings suggest that the specificity of lesion network maps may be more appropriately assessed relative to the co-occurrence of the symptoms studied and to connectome-independent lesion-symptom maps than against the expectation of fully distinct maps.

Several limitations should be considered. Our analyses focused on broad cognitive domains that comprise multiple cognitive processes and frequently co-occur after stroke. Although our analyses indicate that residual similarity between maps largely reflects shared behavioural variance, broad domains limit the anatomical specificity attainable, and finer-grained cognitive measures may yield more distinct networks. Relatedly, cognitive outcomes were based on post-hoc harmonized data and dichotomized into impaired and unimpaired groups rather than analysed as continuous measures, with single-time-point assessment, differential recovery across domains, and domain-specific floor or missing data effects also affecting the anatomical specificity attainable. Finally, our study addresses whether LNM can produce symptom-specific networks; it does not establish the causal role of individual network nodes, nor whether the network-level component improves the prediction of cognitive outcomes beyond lesion location. Moreover, because the domains studied here showed significant lesion-symptom associations in VLSM, our findings do not test the application of LNM to symptoms that cannot be localized from lesion information alone.

In conclusion, our findings indicate that the convergence recently described in lesion network mapping, i.e., the tendency of maps derived from clinically distinct symptoms to resemble one another and to align with generic connectome properties, is not an inherent limitation of the technique but depends critically on study design. Symptom-anchored case-control analyses preserved anatomical specificity with both label permutation-based and parametric inference, with label permutation offering a non-parametric framework that does not rely on independence assumptions. These results identify a practical methodological framework for obtaining biologically meaningful network maps and provide reassurance for the previous LNM studies that already adopted this approach.

## Data availability

The domain-level lesion network maps are provided on Open Science Framework (https://osf.io/puq36). The individual-level data that support the findings of this study are available from the project leads on reasonable request (https://metavcimap.org/group/become-a-member/). Restrictions related to privacy and personal data sharing regulations and informed consent may apply. The code reproducing all analyses and figures is provided on GitHub (https://github.com/umcu-VCI-group/2026_petersen-revisiting_lesion_network_mapping).

## Author contributions

Each author has made a significant contribution to the manuscript and all authors read and approved its final version. M.P.: Conceptualization, Funding, Data curation, Formal analysis, Investigation, Methodology, Project administration, Resources, Software, Visualization, Writing – original draft, Writing – review & editing; K.P.: Investigation, Writing – review & editing; S.B.E.: Investigation, Writing – review & editing; G.J.B.: Conceptualization, Funding, Investigation, Resources, Supervision, Writing – review & editing.

## Acknowledgments

The authors thank all participants of the Meta VCI Map Consortium. The authors also wish to acknowledge the contributions of all researchers who provided resources to the Meta VCI Map post-stroke cognitive impairment initiative: Nick Weaver, PhD; J Matthijs Biesbroek, PhD; Hugo J Kuijf, PhD; Hugo P Aben, MD; Jill Abrigo, MD; Prof Hee-Joon Bae, PhD; Mélanie Barbay, MD; Jonathan G Best, MD; Prof Régis Bordet, PhD; Francesca M Chappell, PhD; Christopher P L H Chen, MD; Prof Martin Dichgans, MD; Thibaut Dondaine, PhD; Jule Filler; Ruben S van der Giessen, PhD; Prof Olivier Godefroy, PhD; Bibek Gyanwali, MD; Olivia K L Hamilton, MSc; Saima Hilal, PhD; Irene M C Huenges Wajer, PhD; Prof Yeonwook Kang, PhD; Prof L Jaap Kappelle, PhD; Beom Joon Kim, PhD; Sebastian Köhler, PhD; Paul L M de Kort, PhD; Prof Peter J Koudstaal, PhD; Gregory Kuchcinski, MD; Bonnie Y K Lam, PhD; Prof Byung-Chul Lee, PhD; Keon-Joo Lee, MD; Jae-Sung Lim, MD; Renaud Lopes, PhD; Stephen D J Makin, PhD; Anne-Marie Mendyk, RN; Prof Vincent C T Mok, MD; Mi Sun Oh, MD; Prof Robert J van Oostenbrugge, PhD; Martine Roussel, PhD; Lin Shi, PhD; Julie Staals, PhD; Maria del C Valdés-Hernández, PhD; Narayanaswamy Venketasubramanian, MBBS; Prof Frans R J Verhey, PhD; Prof Joanna M Wardlaw, PhD; Prof David J Werring, PhD; Xu Xin, PhD; Prof Kyung-Ho Yu, PhD; Martine J E van Zandvoort, PhD; Lei Zhao, PhD. During the preparation of this manuscript, the authors used Claude (Anthropic) to improve language and readability of author-written text.

## Supplementary material

Supplementary material is available at *Brain* online.

