## Supplementary materials for "Ensuring anatomical specificity in lesion network mapping: a multicentre study of 2,950 stroke patients"

**Supplementary Figures**

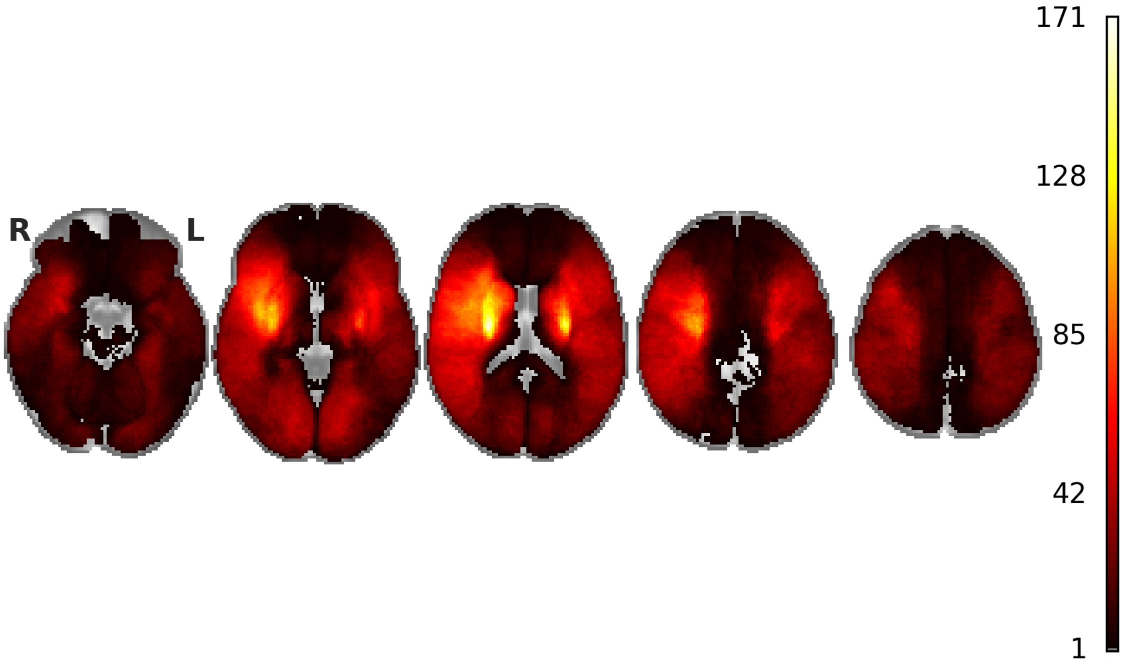


**Supplementary figure S1 | Lesion coverage.** Lesion coverage is shown as a heatmap illustrating the number of stroke lesions per voxel.

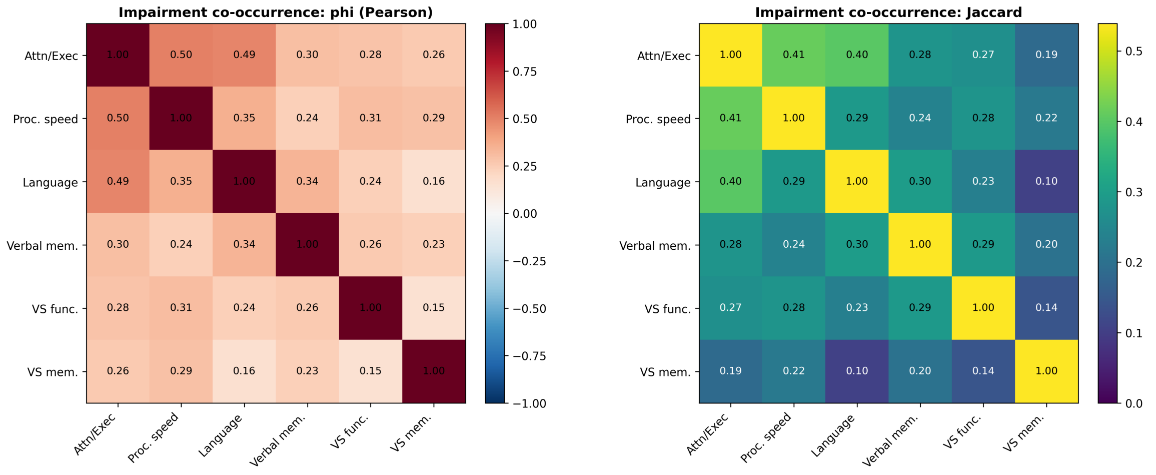


**Supplementary figure S2 | Pairwise co-occurrence of the binary impairment labels across patients.** For each pair of cognitive domains, the heatmaps show how often impairments (impaired vs. unimpaired) coincide in the same patients, quantified as phi coefficient (left) and Jaccard index (right). Abbreviations: Attn/Exec = attention/executive function; Proc. speed = processing speed; Verbal mem. = verbal memory; VS func. = visuospatial functions; VS mem. = visuospatial memory.

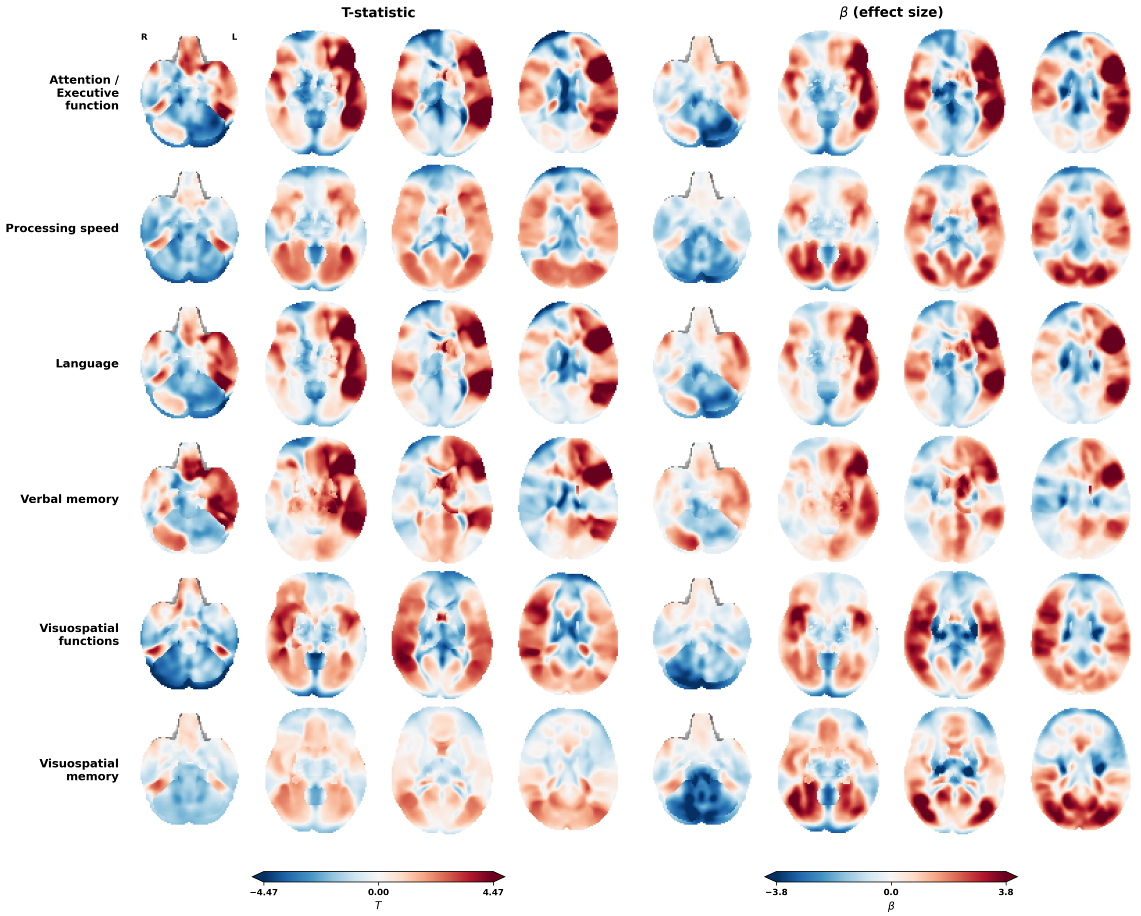


**Supplementary figure S3 | Lesion network maps without significance threshold on axial slices.** Voxelwise statistics of the primary case-control analysis, in which the lesion-derived connectivity maps of cognitively impaired and unimpaired patients were contrasted in a general linear model with cohort as the only covariate. Shown without significance threshold, the maps display the full spatial pattern of the contrast for each of the six cognitive domains: (***a***) T-statistics; (***b***) regression coefficients (β). The colour scale is shared across all domains within each panel (blue, impaired < unimpaired; red, impaired > unimpaired).

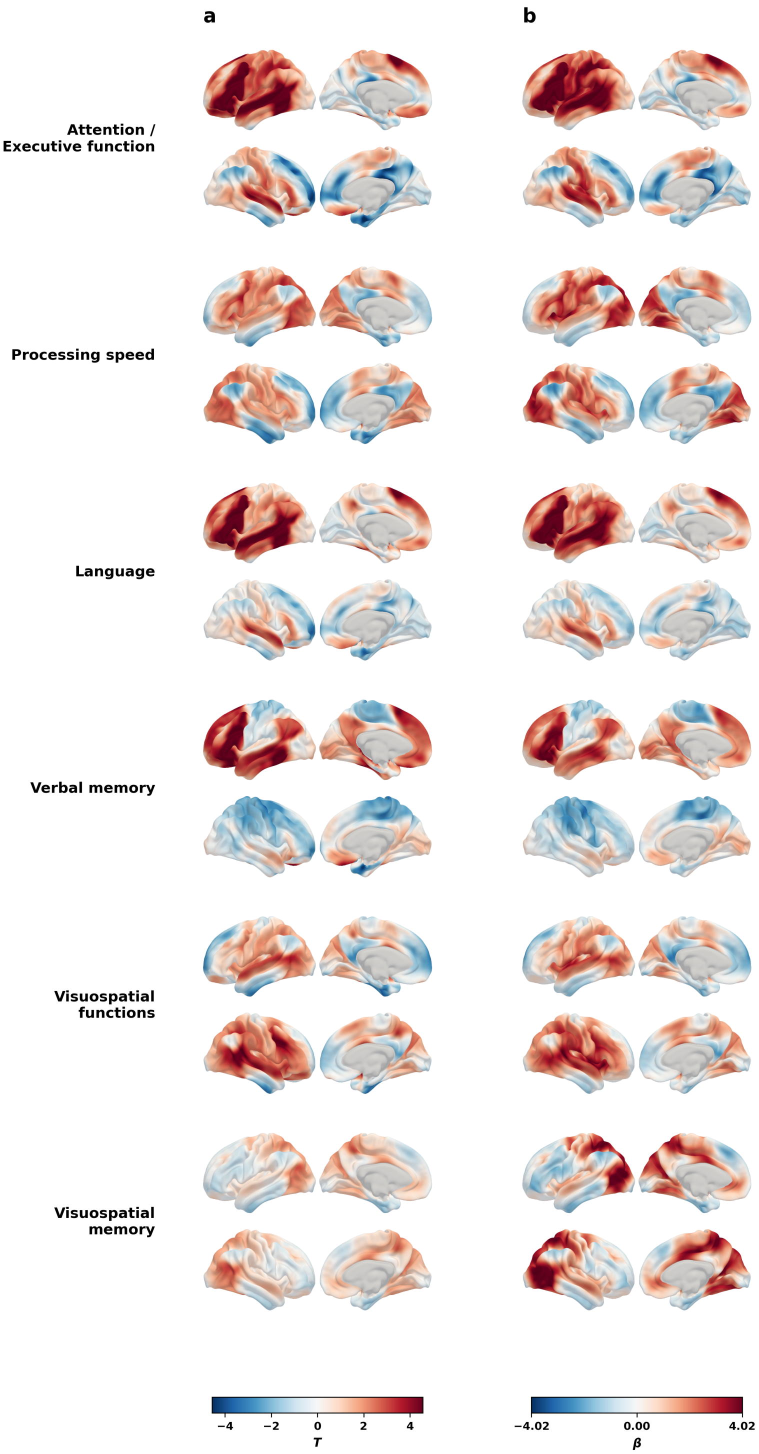


**Supplementary figure S4 | Lesion network maps without significance threshold on cortical surfaces.** Unthresholded lesion network maps for the impaired-versus-unimpaired contrast across the six cognitive domains, projected onto cortical surfaces (lateral and medial views, both hemispheres). (***a***) T-statistics. (***b***) Regression coefficients (β). No significance threshold is applied; the colour scale is shared across all domains within each panel (blue, impaired < unimpaired; red, impaired > unimpaired).

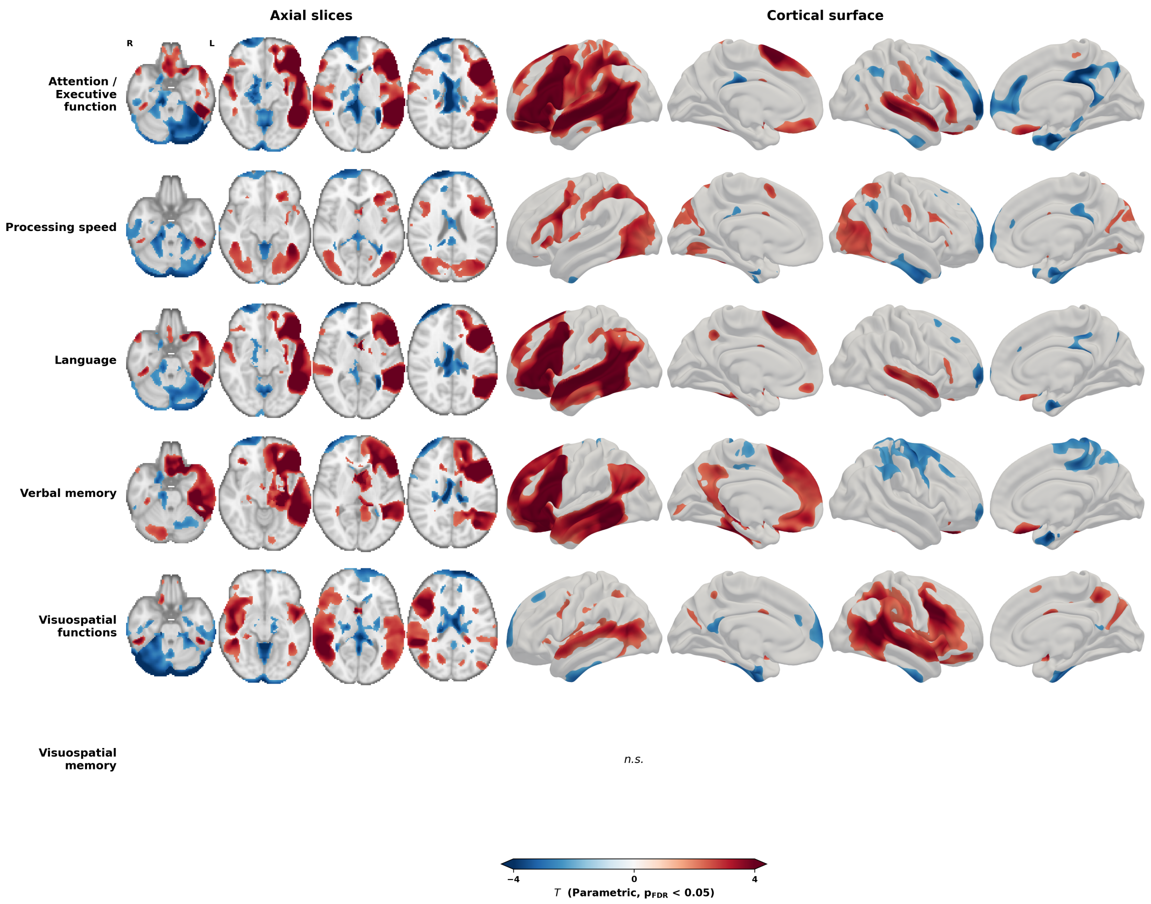


**Supplementary figure S5 | Parametric lesion network maps on axial slices.** As a second case-control approach, impaired and unimpaired patients were compared with a parametric two-sample t-test, using the same general linear model as the label permutation-based analysis (cohort as the only covariate) but deriving p-values from the t-distribution rather than from symptom-label permutation. Maps for the six cognitive domains are thresholded at $p_{FDR} < 0.05$ and shown on four axial slices (z = −26, −10, 6, 22 mm; radiological orientation) as well as cortical surface renderings (Conte69 midthickness); blue, impaired < unimpaired; red, impaired > unimpaired. No voxels survived for visuospatial memory (n.s.). The colour scale is shared across panels.

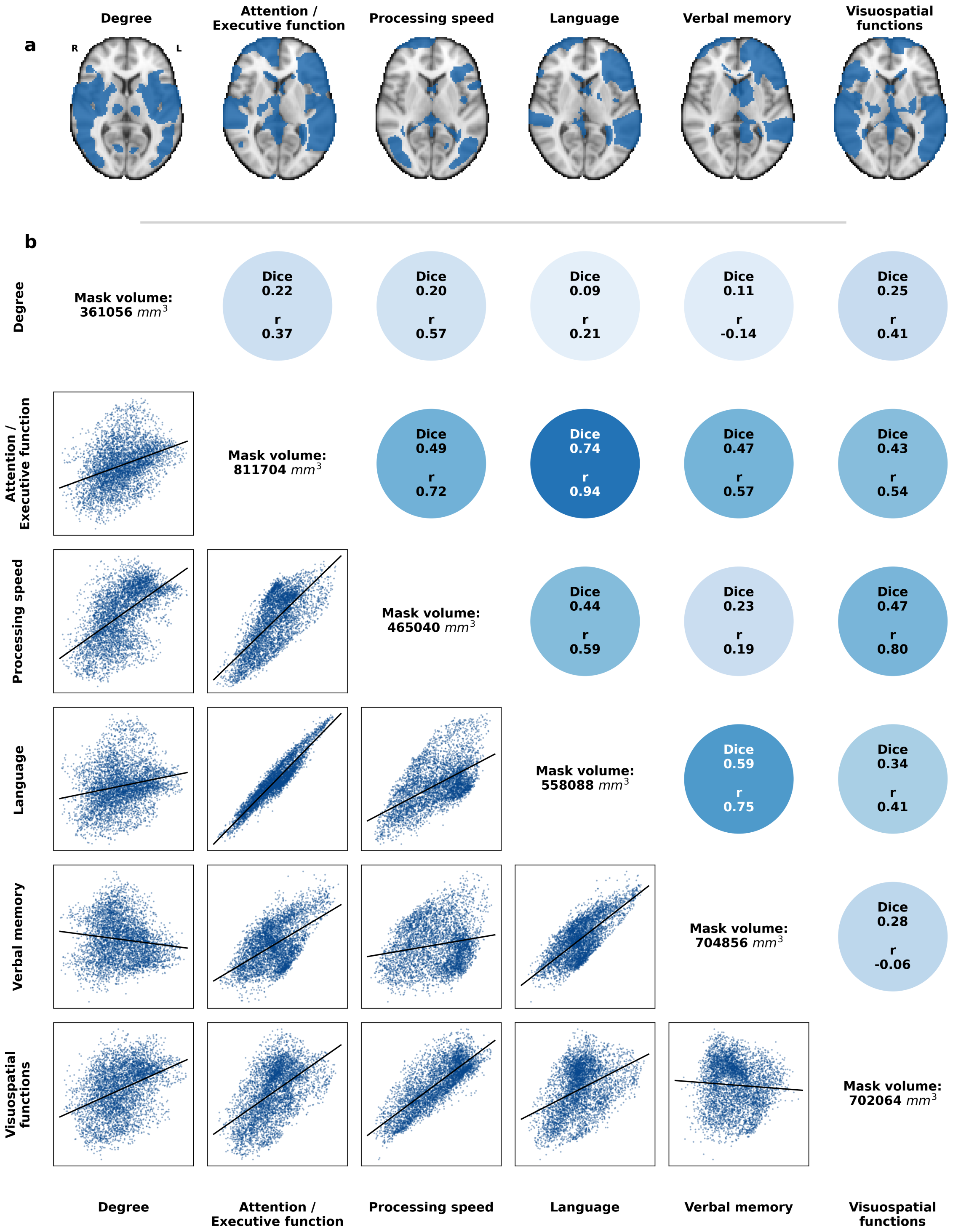


**Supplementary figure S6 | Spatial correspondence of parametric lesion network maps.** To test whether cross-domain similarity and alignment with the normative degree map depend on the inference approach, the spatial correspondence analysis presented in the main manuscript for the label permutation-based maps was repeated for the parametric maps. (a) Symptom-linked lesion network maps derived using parametric two-sample t-tests, thresholded at $p_{FDR} < 0.05$. Visuospatial memory is not shown, as no voxels survived thresholding. (b) Pairwise comparisons of lesion network maps. Upper triangle: similarity quantified as Dice overlap of significance-thresholded maps. The first row/column compares each map with the normative connectome degree map (degree defined as the row-sum of the normative FC matrix; binary degree mask thresholded at the 95th percentile). Lower triangle: pairwise spatial correlation illustrated by regression plot of voxels from the unthresholded T-statistic maps. Diagonal: volumes of thresholded masks of degree and significance maps ($mm^3$).

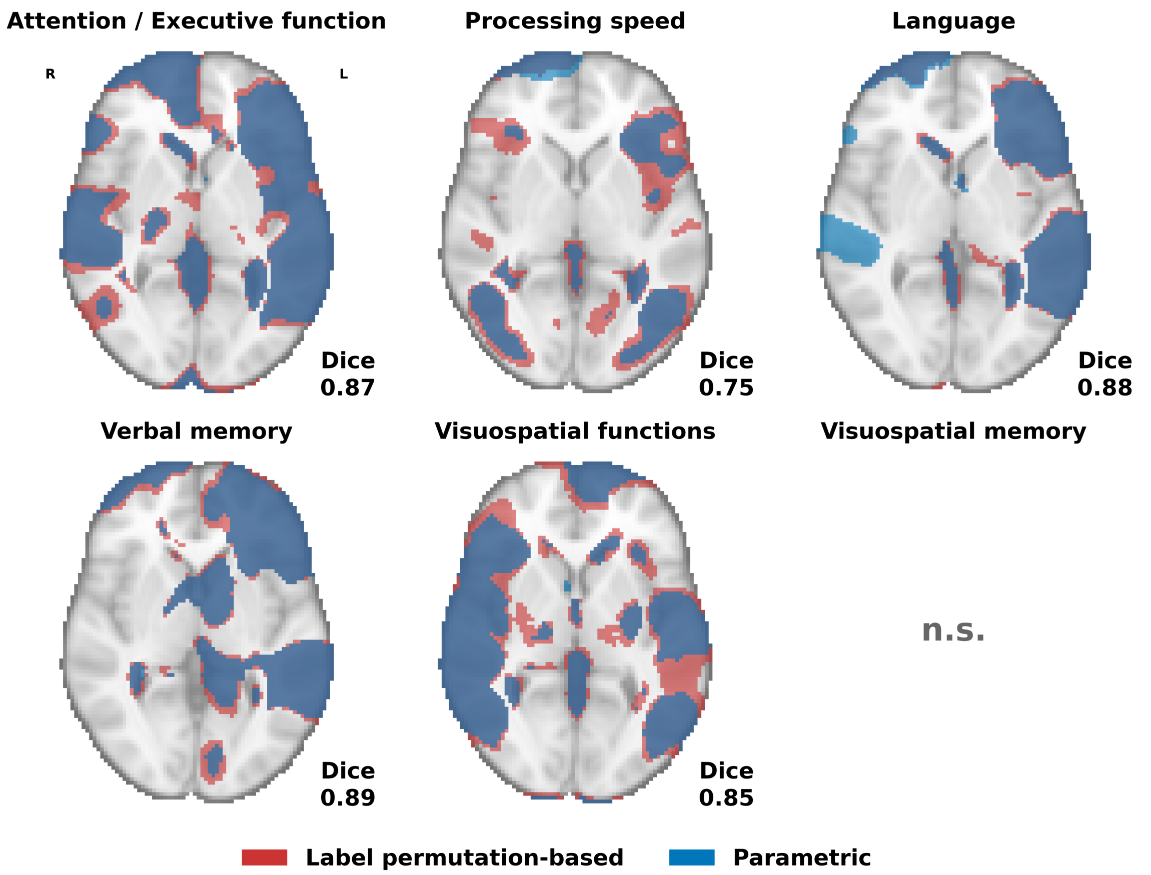


**Supplementary figure S7 | Comparison of label permutation-based and parametric inference in lesion network mapping.** To test whether the inference approach changes which voxels are deemed significant, the label permutation-based (red) and parametric (blue) maps of each cognitive domain, both thresholded at $p_{FDR} < 0.05$, are overlaid; overlapping areas appear dark blue. Dice coefficients quantify the overlap for each domain.

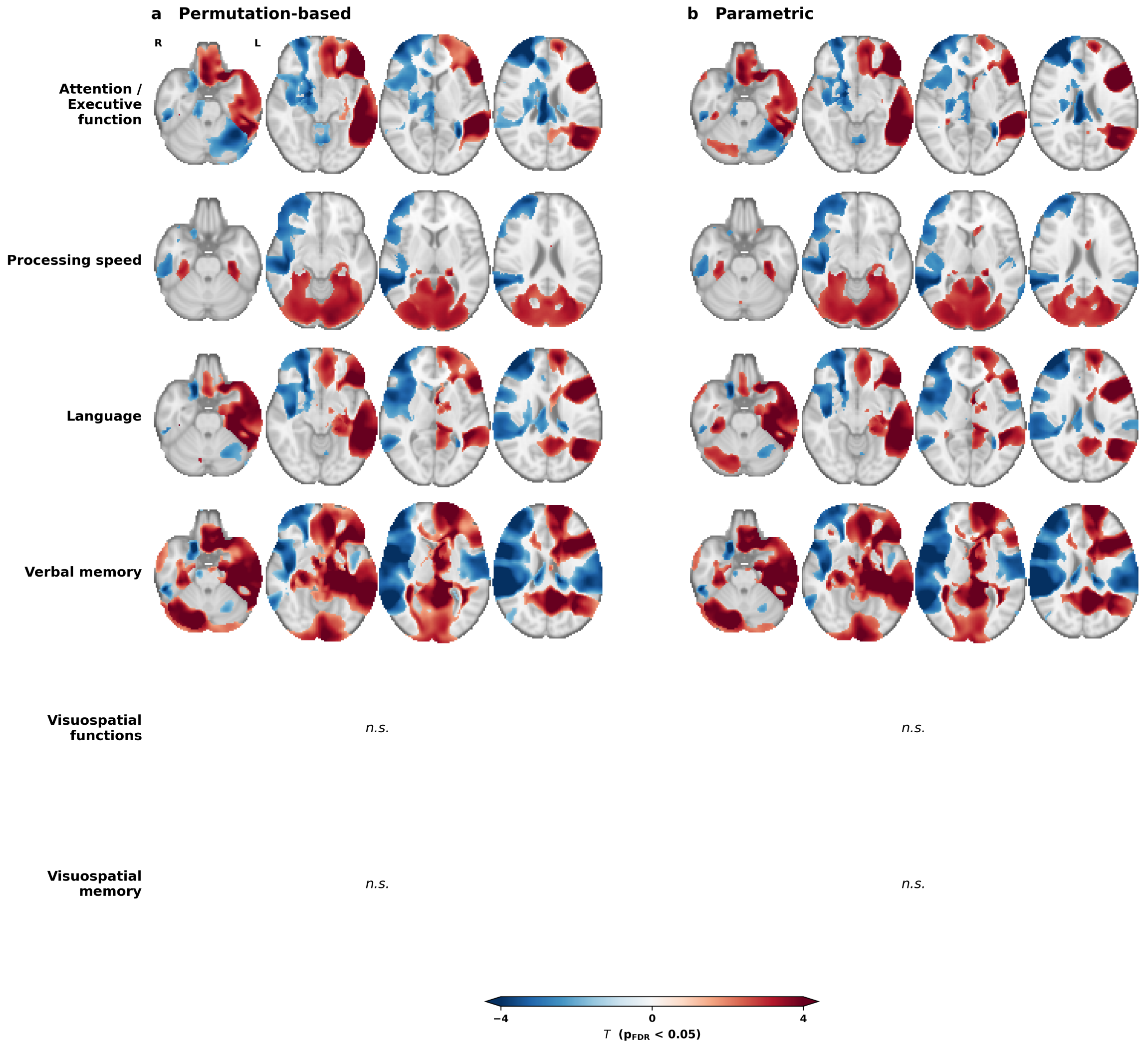


**Supplementary figure S8 | Label permutation-based and parametric lesion network maps after adjusting for lesion volume and NIHSS.** To test whether the case-control maps are driven by lesion size or stroke severity, both case-control analyses were repeated with lesion volume and National Institutes of Health Stroke Scale (NIHSS) score as additional covariates. Maps for the six cognitive domains are thresholded at $p_{FDR} < 0.05$ and shown on four axial slices (z = −26, −10, 6, 22 mm; radiological orientation); blue, impaired < unimpaired; red, impaired > unimpaired. (**a**) Label permutation-based, (**b**) parametric. No voxels survived for visuospatial functions and visuospatial memory (n.s.). The colour scale is shared across panels.

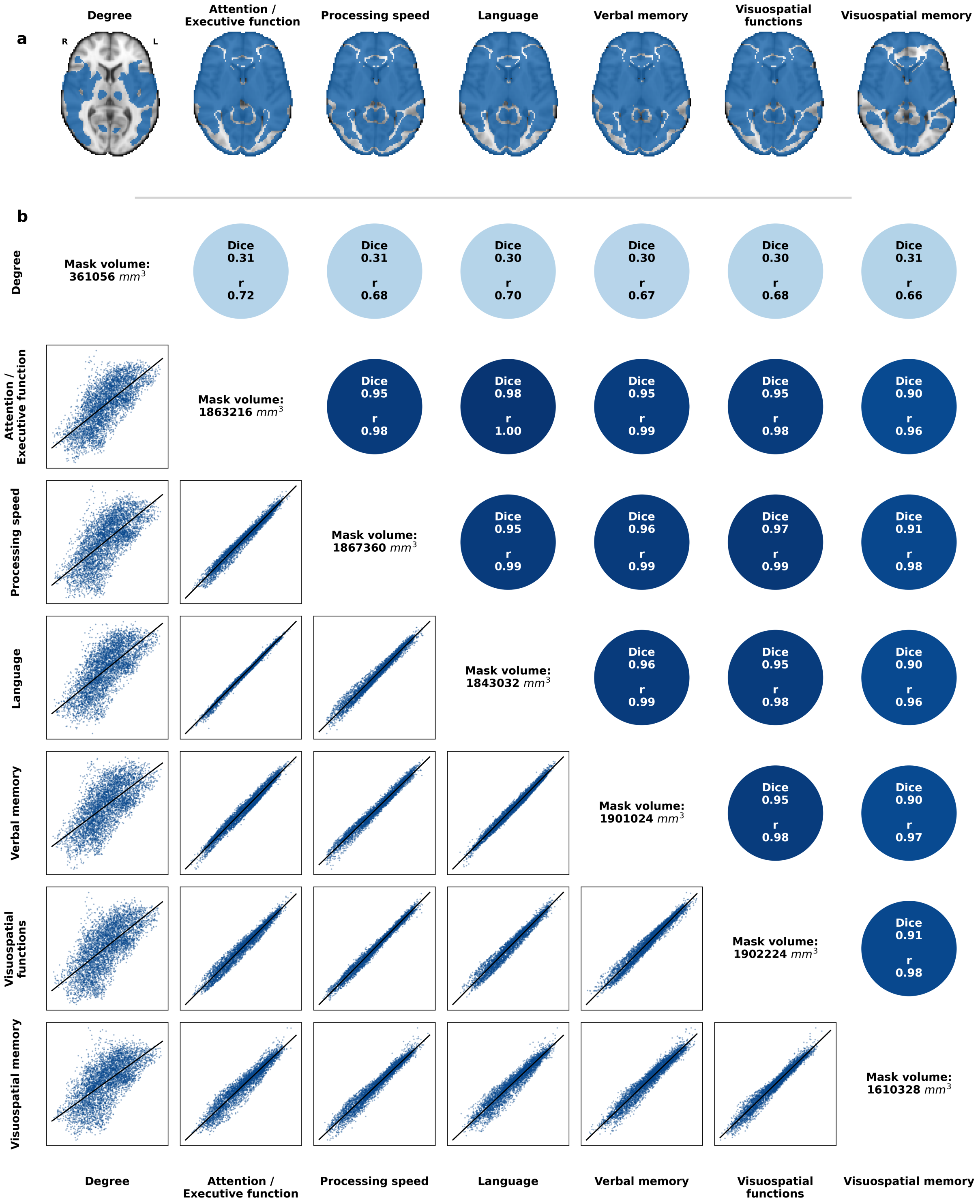


**Supplementary figure S9 | Spatial correspondence of one-sample t-test results.** (a) Lesion network maps derived using one-sample t-tests in cognitively impaired patients, thresholded at $p_{FDR} < 0.05$. (b) Pairwise comparisons of lesion network maps from one-sample t-tests. Upper triangle: similarity quantified as Dice overlap of significance-thresholded maps. The first row/column compares each map with the normative connectome degree map (degree defined as the row-sum of the normative FC matrix; binary degree mask thresholded at the 95th percentile). Lower triangle: pairwise spatial correlation illustrated by regression plot of voxels from the unthresholded T-statistic maps. Diagonal: volumes of thresholded masks of degree and significance maps ($mm^3$).

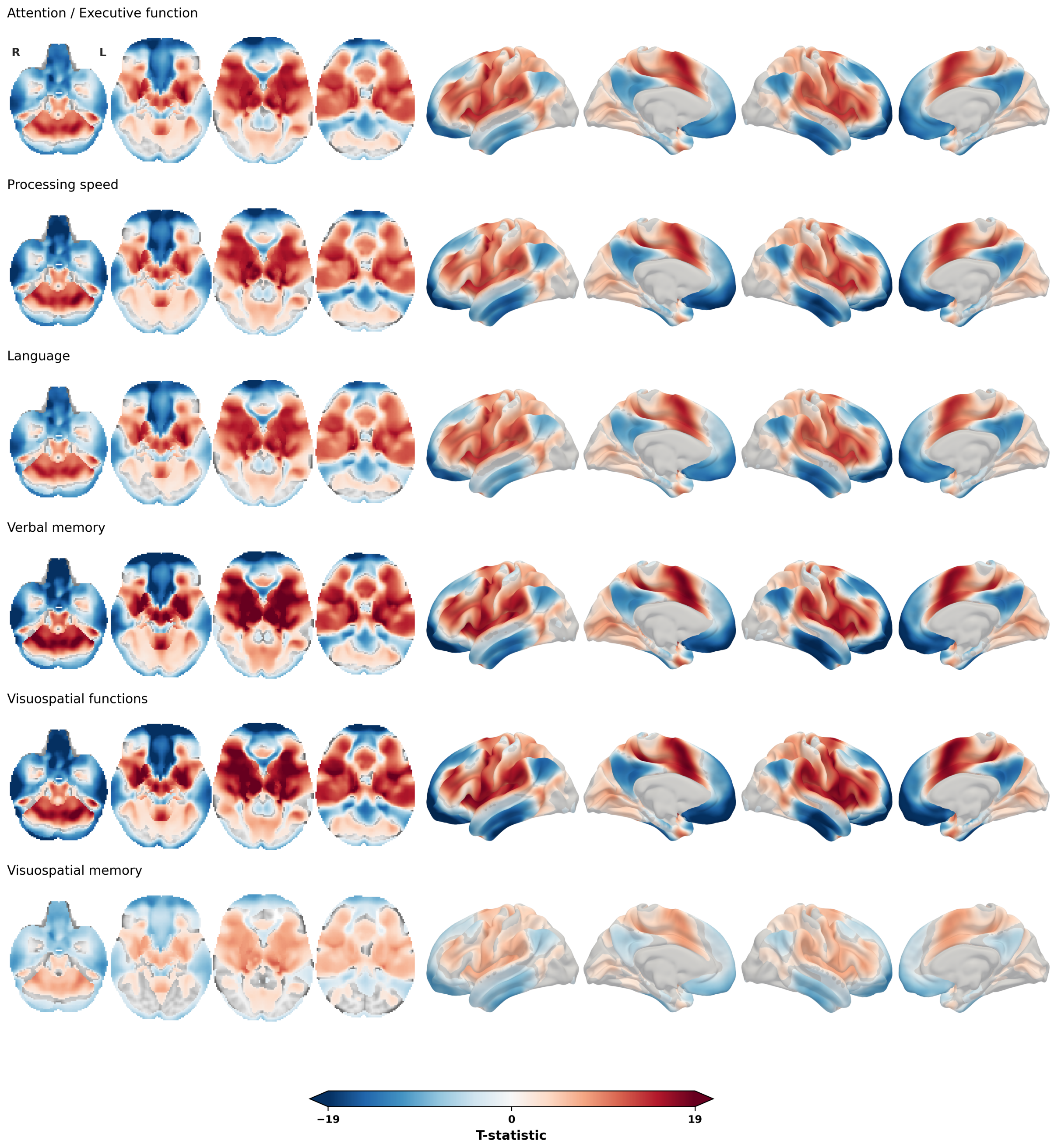


**Supplementary figure S10 | One-sample t-test results.** Lesion network maps across the six cognitive domains based on one-sample t-tests of cognitively impaired patients, thresholded at $p_{FDR} < 0.05$ and shown on four axial slices (z = −26, −10, 6, 22 mm; radiological orientation) as well as cortical surface renderings (Conte69 midthickness); red, voxels positively correlated with the lesions of impaired patients; blue, voxels anticorrelated. One-sample t-tests were adjusted for cohort. The colour scale is shared across panels.

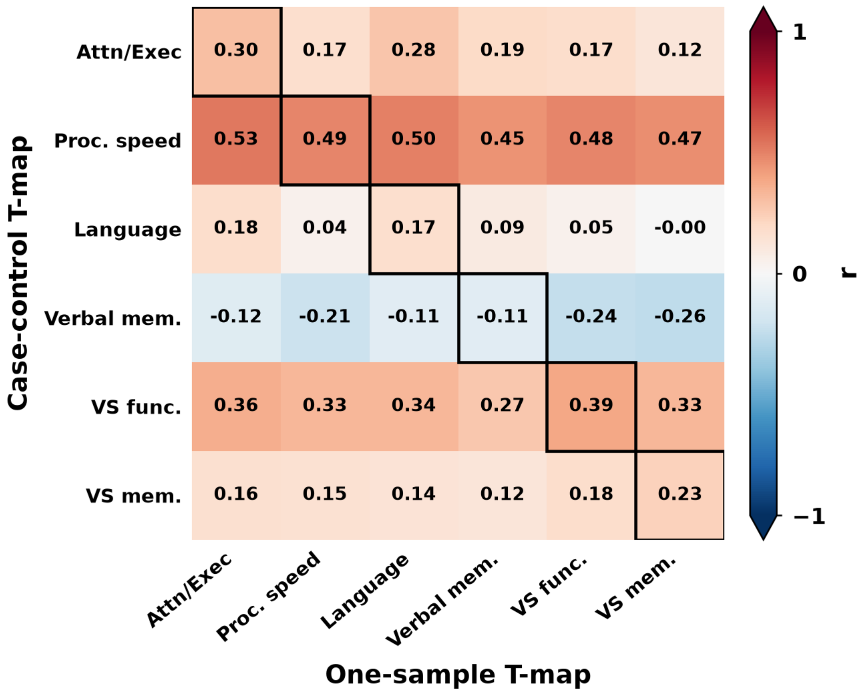


**Supplementary figure S11 | Correspondence between case-control maps and one-sample t-test maps.** To test whether the one-sample and case-control designs identify the same domain-specific patterns, the heatmap shows voxelwise Pearson correlations between the unthresholded case-control T-maps (rows) and one-sample T-maps (columns) for all pairs of domains. Black outlines mark the own-domain comparisons (diagonal), where domain-specific correspondence would appear as higher values. Own-domain correspondence was low ($r = -0.11$ to $0.49$), and each case-control map correlated near-uniformly with all six one-sample maps. All models were adjusted for cohort.

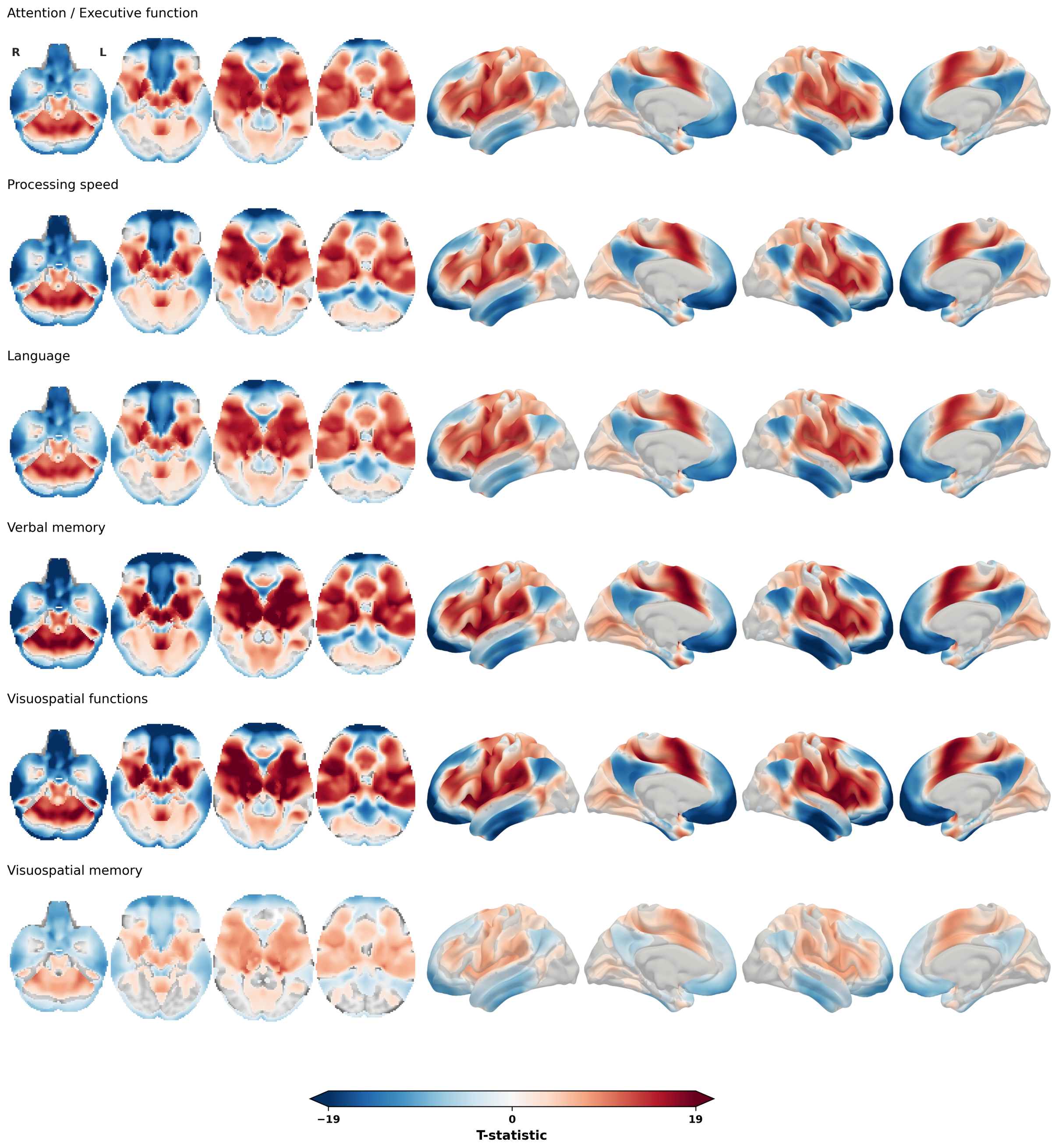


**Supplementary figure S12 | One-sample t-test lesion network maps adjusted for cohort, lesion volume, and NIHSS.** Fully covariate-adjusted one-sample analyses, thresholded at $p_{FDR} < 0.05$ (TFCE). The maps matched those resulting from the one-sample t-tests only adjusted for cohort (voxelwise $r \geq 0.99$) and retained the degree map convergence.

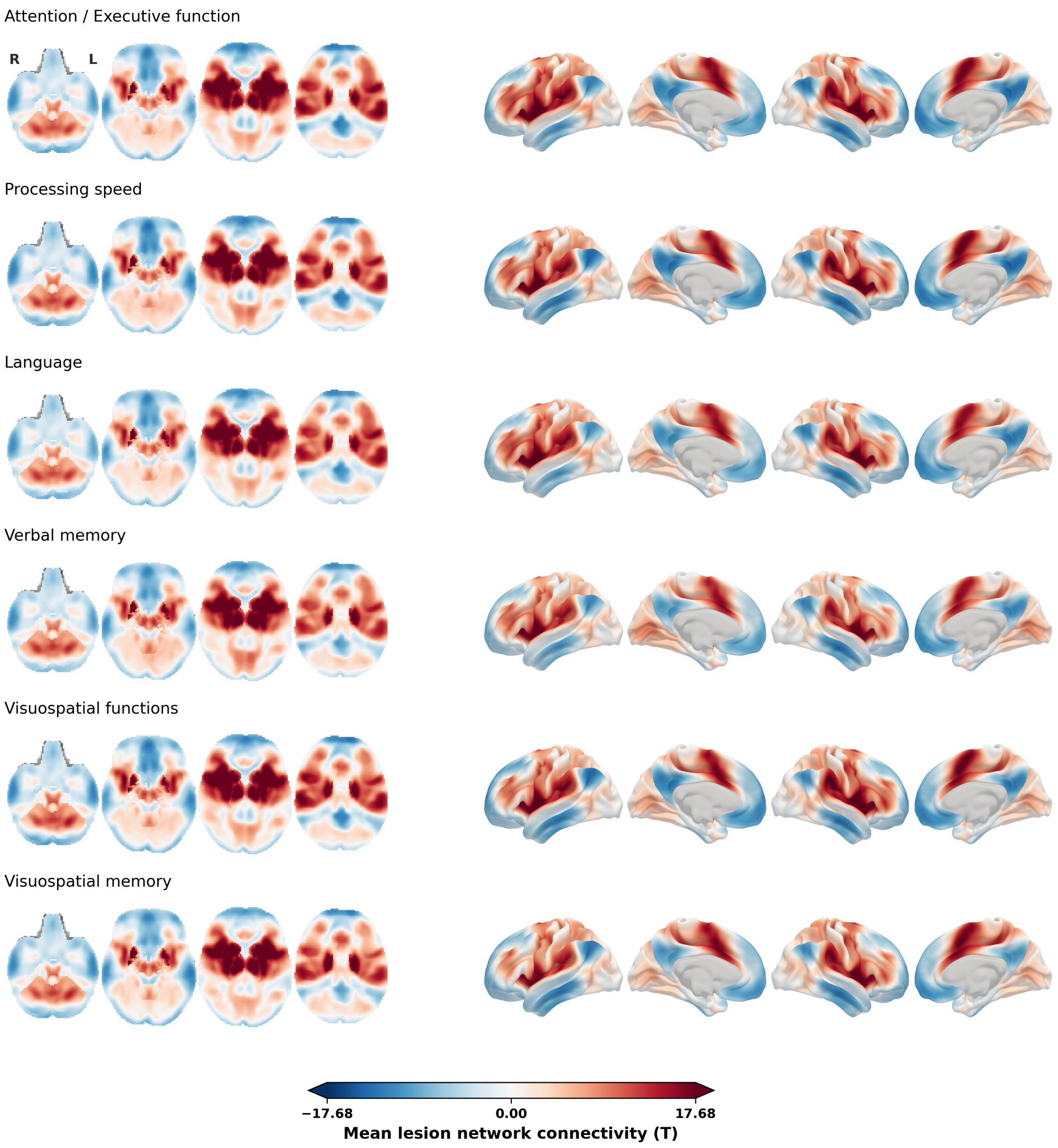


**Supplementary figure S13 | Mean aggregation maps of cognitively impaired patients.** Voxelwise mean of the lesion-derived connectivity maps across cognitively impaired patients per cognitive domain which is the same aggregation procedure examined in the previous critique. The mean maps aligned closely with the maps resulting from one-sample t-tests (voxelwise r = 0.91–0.93 per domain) and correlated similarly strongly with the normative degree map (r = 0.72–0.77; concordant paired comparison against the case-control maps: t(5) = 6.28, p = 0.002), demonstrating that averaging and one-sample t-testing behave interchangeably.

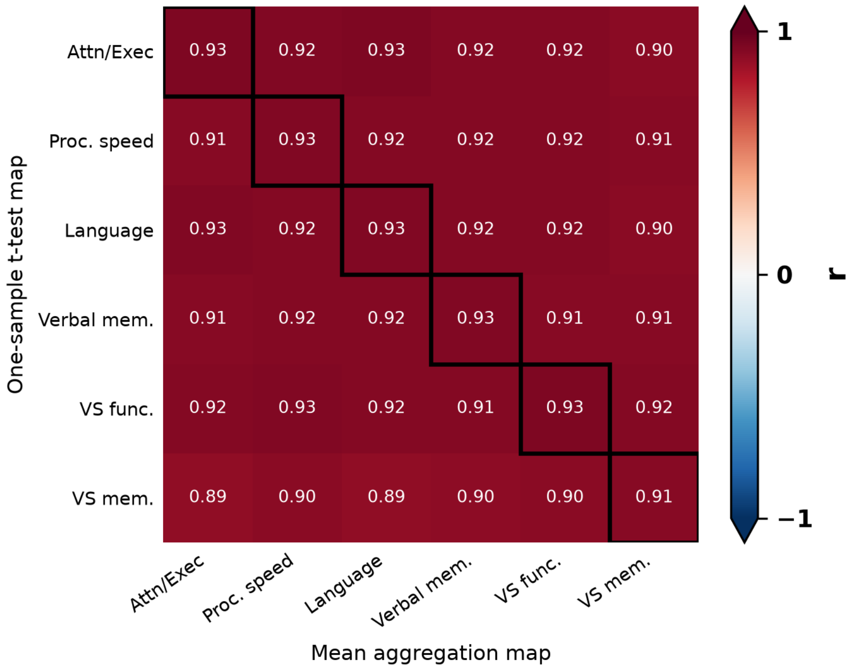


**Supplementary figure S14 | Correspondence between one-sample t-test and mean aggregation maps.** Voxelwise Pearson correlations between the one-sample t-test maps (rows) and the complementary mean aggregation maps (columns) across the six cognitive domains (in-brain voxels); black outlines mark the own-domain comparisons (diagonal). Own-domain correspondence was very high (r = 0.91–0.93), and all cross-domain cells were similarly high (r = 0.89–0.93), reflecting the near-identity of both aggregation procedures and their convergence across domains: the T-statistic is the voxelwise mean scaled by its standard error, so the two maps differ only by the voxelwise variability of the connectivity maps.

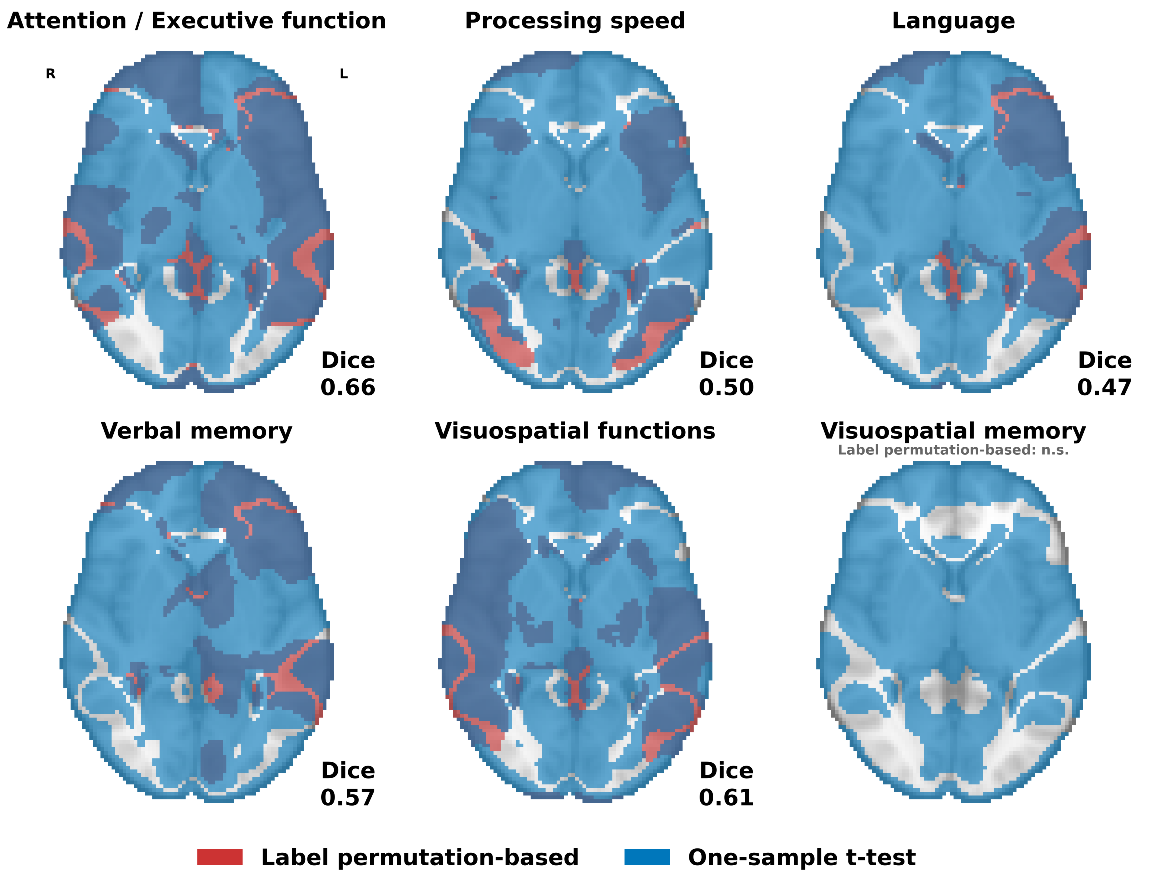


**Supplementary figure S15 | Comparison of label permutation-based inference and one-sample t-test in lesion network mapping.** For each cognitive domain, lesion network maps thresholded at $p_{FDR} < 0.05$ are shown for label permutation-based inference (red) overlaid with the one-sample t-test maps of cognitively impaired patients (blue); overlapping areas appear dark blue. Both inference approaches were adjusted for cohort. Dice coefficients quantify overlap (mean Dice = 0.56 ± 0.07 across the five domains with significant label permutation-based maps; no significant voxels for visuospatial memory).

n.s. = no significant voxels.

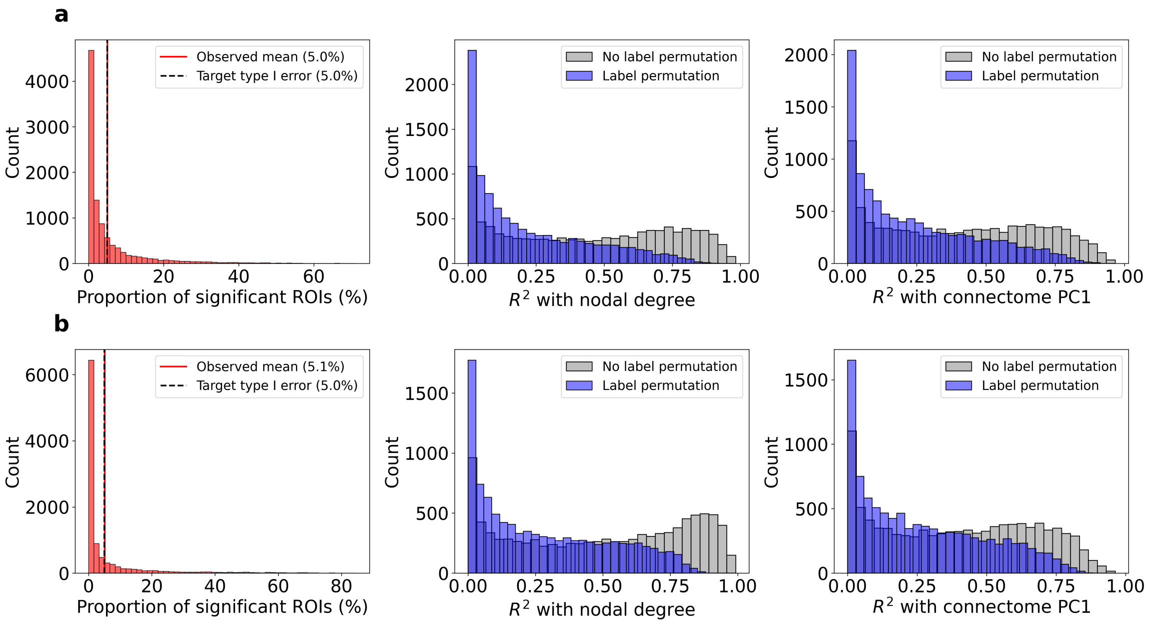


**Supplementary figure S16 | Simulation-based validation of type I error control under symptom-label permutation using real and synthetic lesions.**

To verify that symptom-label permutation controls false positives while accounting for nonspecific connectome structure, null studies without any true lesion–symptom relationship were simulated.

Simulation design was informed by prior simulation studies by van den Heuvel et al., 2026 (described in their supplementary text S9 – Simulation; https://doi.org/10.1038/s41593-025-02196-7). For computational efficiency, simulations were conducted at the ROI level rather than the voxel-level analyses used for empirical data in the main manuscript.

Simulations used ROI-level lesion masks and a normative functional connectivity matrix derived from the GSP1000 dataset, based on the Schaefer1000 cortical atlas and the Tian54 subcortical atlas (1,054 regions/nodes).

Lesions were generated using two approaches, analysed separately:

1. **Synthetic lesions.** For each of 10,000 simulation runs, 50 synthetic lesions were generated. Each lesion consisted of a randomly selected set of lesioned ROIs, with lesion size (number of ROIs) sampled uniformly between 1 and 50.
2. **Real lesions.** For each of 10,000 simulation runs, 50 lesions were randomly sampled with replacement from the 2,950 Meta VCI Map patients.

For each simulation run, lesion vectors were concatenated into a lesion matrix $\mathrm{M}$ (patients × ROIs). Symptom-linked lesion network maps were then computed using the simplified symptom-linked lesion network mapping formulation sLNM $= \mathrm{sv} \times (\mathrm{M} \times \mathrm{C})$, where $\mathrm{C}$ denotes the normative functional connectivity matrix and $\mathrm{sv}$ is a standardized symptom vector.

To ensure the absence of a true lesion–symptom relationship, symptom scores $sv$ were drawn from a standard normal distribution independently of the lesion data. The matrix product $\mathrm{M} \times \mathrm{C}$ yields the functional connectivity profiles associated with each lesion, after which these profiles were linked across lesions with the symptom vector $sv$ to obtain the resulting sLNM map.

Symptom–label permutation was then performed (1,000 permutations per simulation run) following the same computational pipeline to generate permuted ROI-level lesion network maps. For each ROI, a permutation p-value was computed as the proportion of permuted statistics greater than or equal to the observed statistic ($|t| \geq |t\_obs|$), and regions were thresholded at $p < 0.05$ (two-tailed) to estimate the empirical false positive rate.

Panels follow identical analysis logic, differing only in lesion generation: **(a)** synthetic lesions and **(b)** real lesions.

**Left:** Distribution of false positive rates (FPR; proportion of ROIs with $p < 0.05$) across 10,000 simulations. The observed mean FPR (red line) matched the nominal type I error rate of 5% (dashed line), indicating appropriate statistical control for both real and synthetic lesions.

**Middle:** Distributions of $R^2$ between lesion network maps and nodal degree for uncorrected raw maps (grey) and permutation-corrected maps (blue) across simulations.

**Right:** Distributions of $R^2$ between lesion network maps and the first principal component (PC1) of the connectome for raw maps (grey) and permutation-corrected maps (blue).

Symptom–label permutation markedly reduced spurious alignment with both nodal degree and connectome PC1, demonstrating that the permutation null model effectively accounts for nonspecific connectome structure. This pattern was observed for both real and synthetic lesions.

Abbreviations: FPR = false positive rate; PC1 = first principal component; ROI = region of interest.

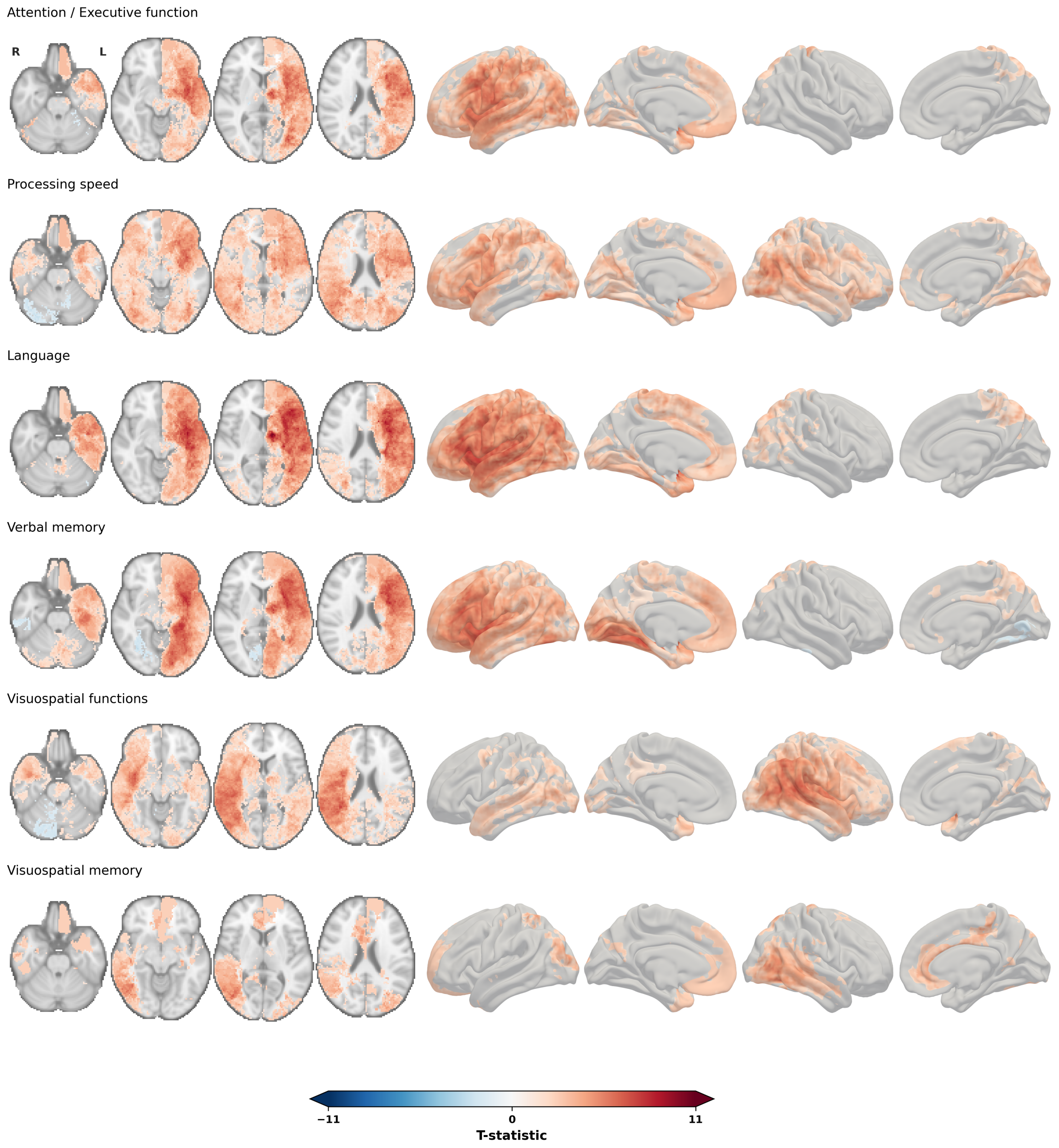


**Supplementary figure S17 | Voxel-based lesion-symptom mapping results.** Lesion-symptom maps across the six cognitive domains, thresholded at $p_{FDR} < 0.05$ and shown on four axial slices (z = −26, −10, 6, 22 mm; radiological orientation) as well as cortical surface renderings (Conte69 midthickness); red, voxels more frequently lesioned in impaired than unimpaired patients; blue, the converse. The colour scale is shared across panels. Models were matched to the primary lesion network mapping analysis (label permutation-based inference; cohort as the only covariate).

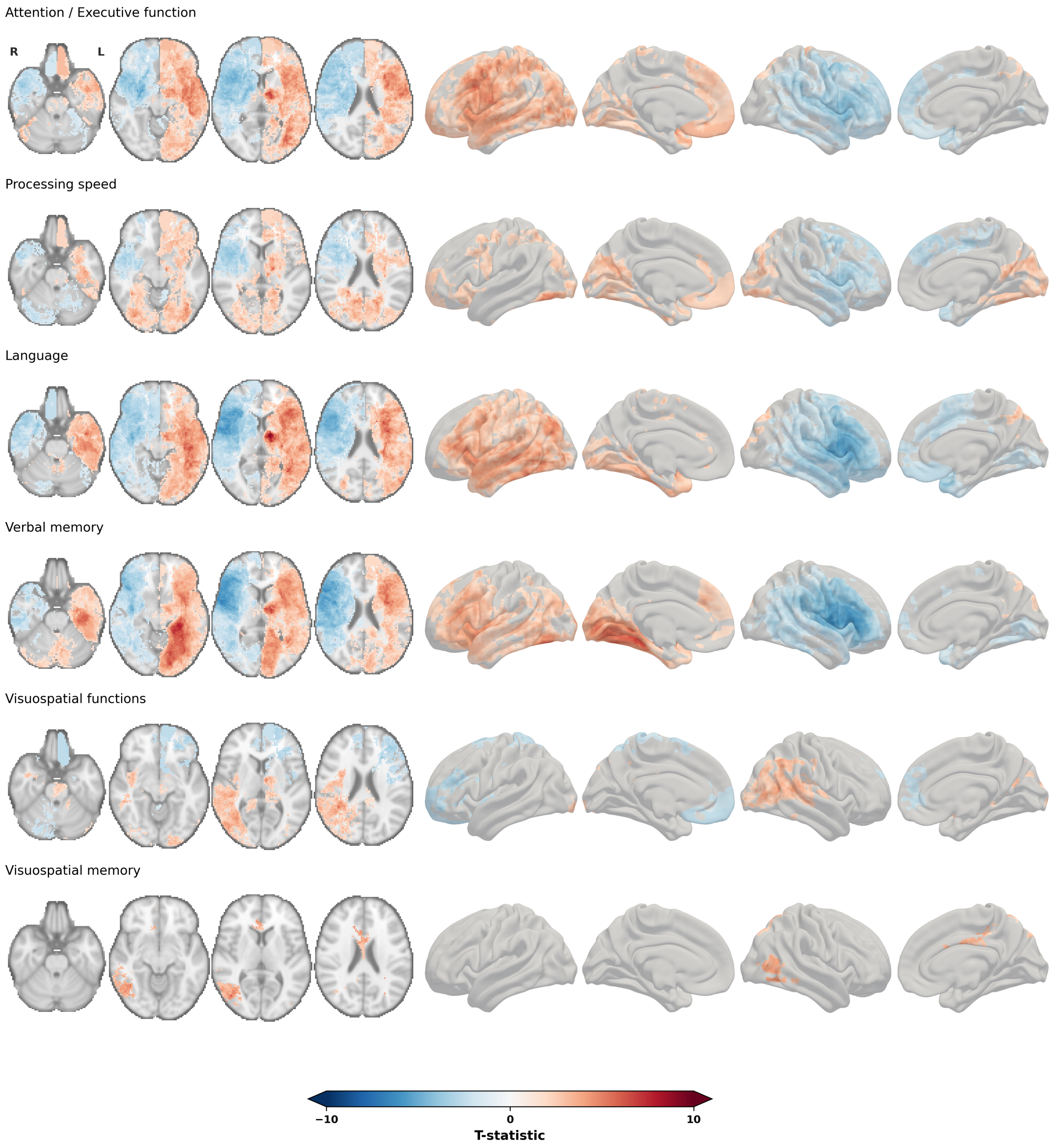


**Supplementary figure S18 | Voxel-based lesion-symptom mapping results, fully adjusted models.** Lesion-symptom maps across the six cognitive domains, adjusted for cohort, lesion volume, and NIHSS (covariate-matched to the fully adjusted lesion network mapping analyses), thresholded at $p_{FDR} < 0.05$ and shown on four axial slices (radiological orientation) and cortical surface renderings; red, voxels more frequently lesioned in impaired than unimpaired patients; blue, the converse. The colour scale is shared across panels.

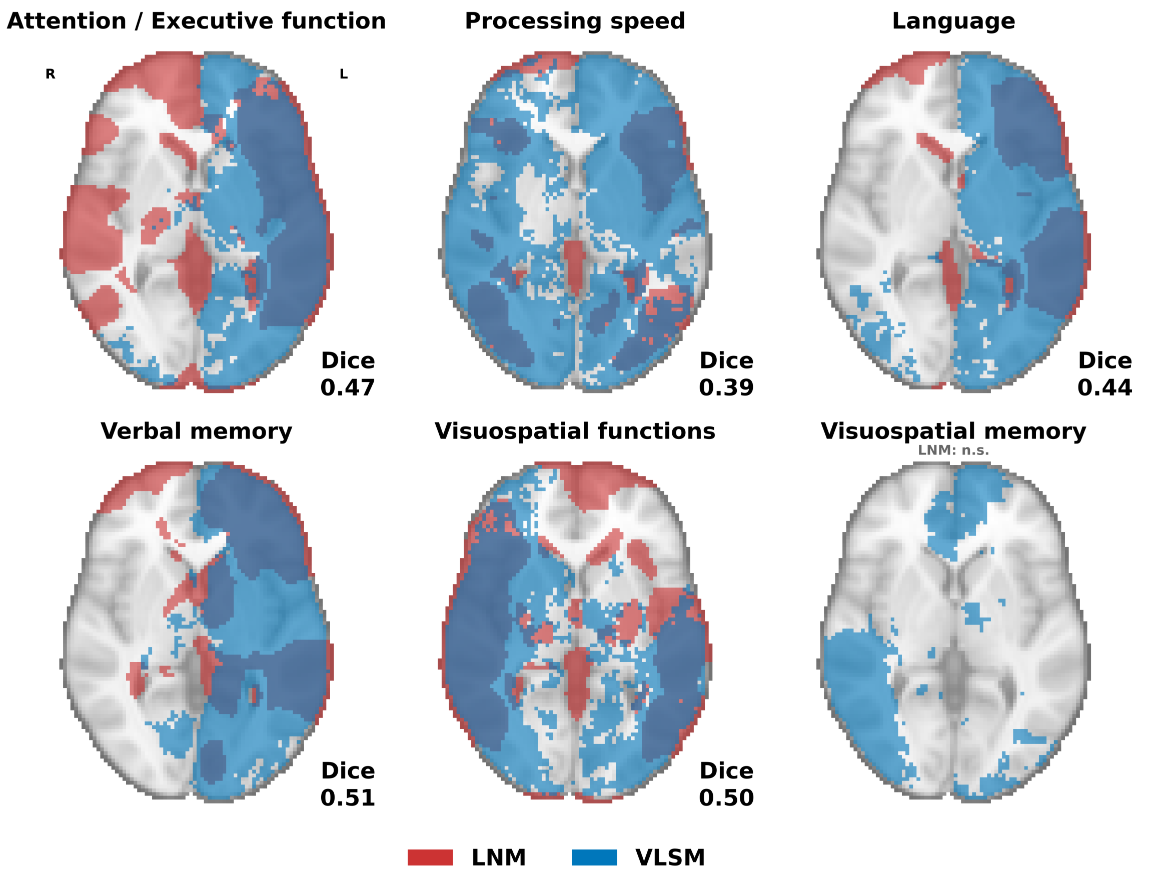


**Supplementary figure S19 | Comparison of label permutation-based lesion network mapping and voxel-based lesion-symptom mapping, adjusted for cohort.** For each cognitive domain, lesion network maps thresholded at $p_{FDR} < 0.05$ (red) are overlaid with the voxel-based lesion-symptom maps (blue; cohort as the only covariate in both analyses); overlapping areas appear dark blue. Dice coefficients quantify overlap (mean Dice = 0.46 ± 0.04 across the five domains with significant lesion network maps).

n.s. = no significant voxels.

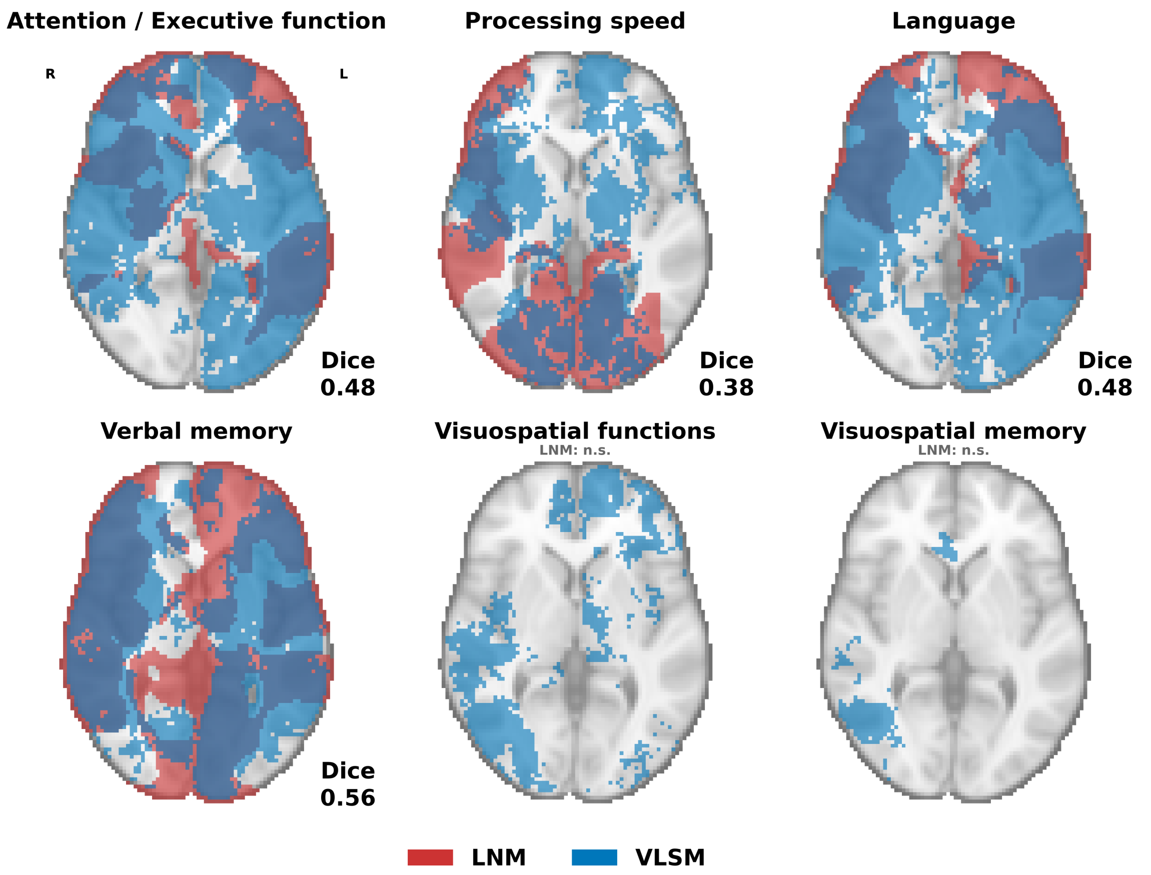


**Supplementary figure S20 | Comparison of label permutation-based lesion network mapping and voxel-based lesion-symptom mapping, fully adjusted models.** For each cognitive domain, lesion network maps thresholded at $p_{FDR} < 0.05$ (red) are overlaid with the voxel-based lesion-symptom maps (blue); both analyses adjusted for cohort, lesion volume, and NIHSS; overlapping areas appear dark blue. Dice coefficients quantify overlap (mean Dice = 0.48 ± 0.07 across the four domains with significant lesion network maps).

n.s. = no significant voxels.

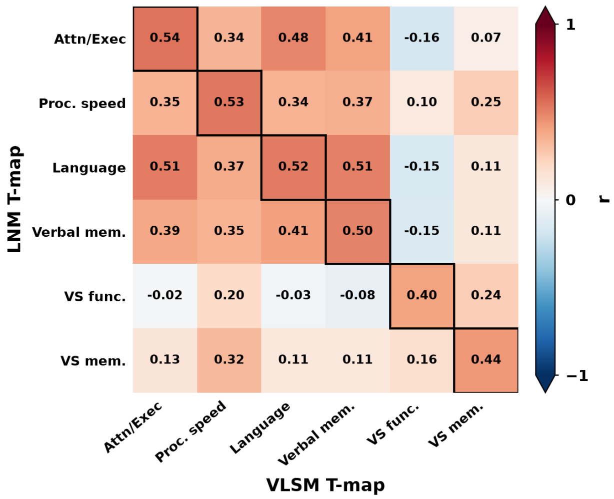


**Supplementary figure S21 | Correspondence between lesion network maps and voxel-based lesion-symptom maps, fully adjusted models.** To test whether the correspondence between both methods is domain-specific, the heatmap shows voxelwise Pearson correlations between the unthresholded lesion network T-maps (rows) and voxel-based lesion-symptom T-maps (columns) for all pairs of domains (in-brain voxels); black outlines mark the own-domain comparisons (diagonal). Both analyses were adjusted for cohort, lesion volume, and NIHSS. Own-domain correlations ranged from r = 0.40 to 0.54 (mean r = 0.49), indicating moderate correspondence between the two methods.

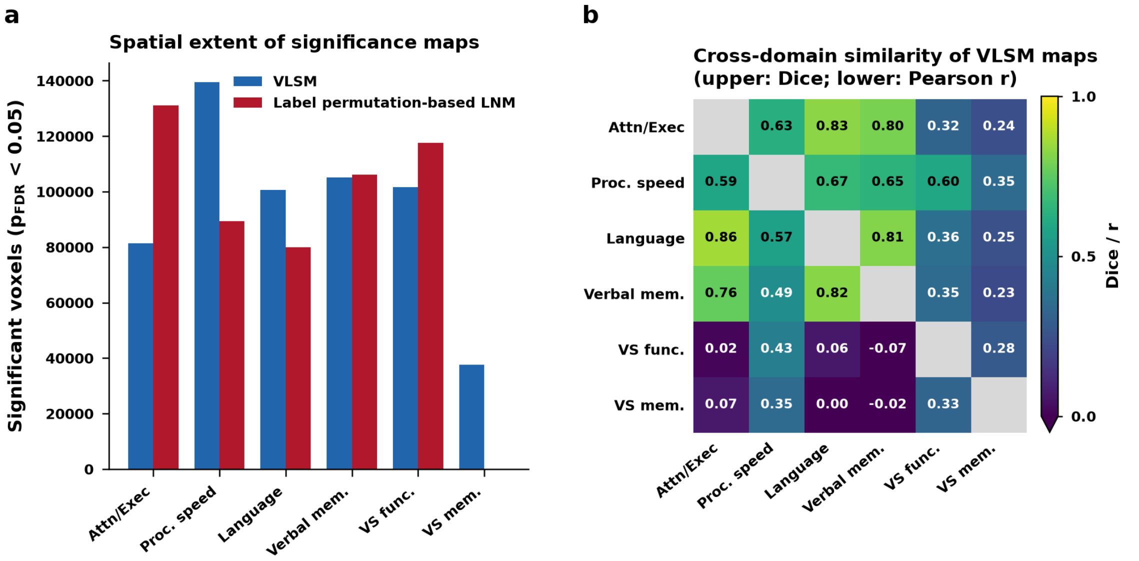


**Supplementary figure S22 | Concordance of voxel-based lesion-symptom maps and label permutation-based lesion network maps.** (a) Spatial extent of the significance maps for each cognitive domain, expressed as the number of significant voxels ($p_{FDR}$ < 0.05), for voxel-based lesion-symptom maps (blue) and label permutation-based lesion network maps (red). The lesion network map for visuospatial memory contained no significant voxels. (b) Cross-domain similarity of the voxel-based lesion-symptom maps: upper triangle, Dice overlap of the significance-thresholded maps ($p_{FDR}$ < 0.05); lower triangle, Pearson correlation of the unthresholded T-maps (in-brain voxels); grey diagonal, self-comparisons. Mean cross-domain Dice = 0.49 ± 0.22; mean Pearson r = 0.35 ± 0.32. Abbreviations: Attn/Exec = attention/executive function; Proc. speed = processing speed; Verbal mem. = verbal memory; VS func. = visuospatial functions; VS mem. = visuospatial memory.

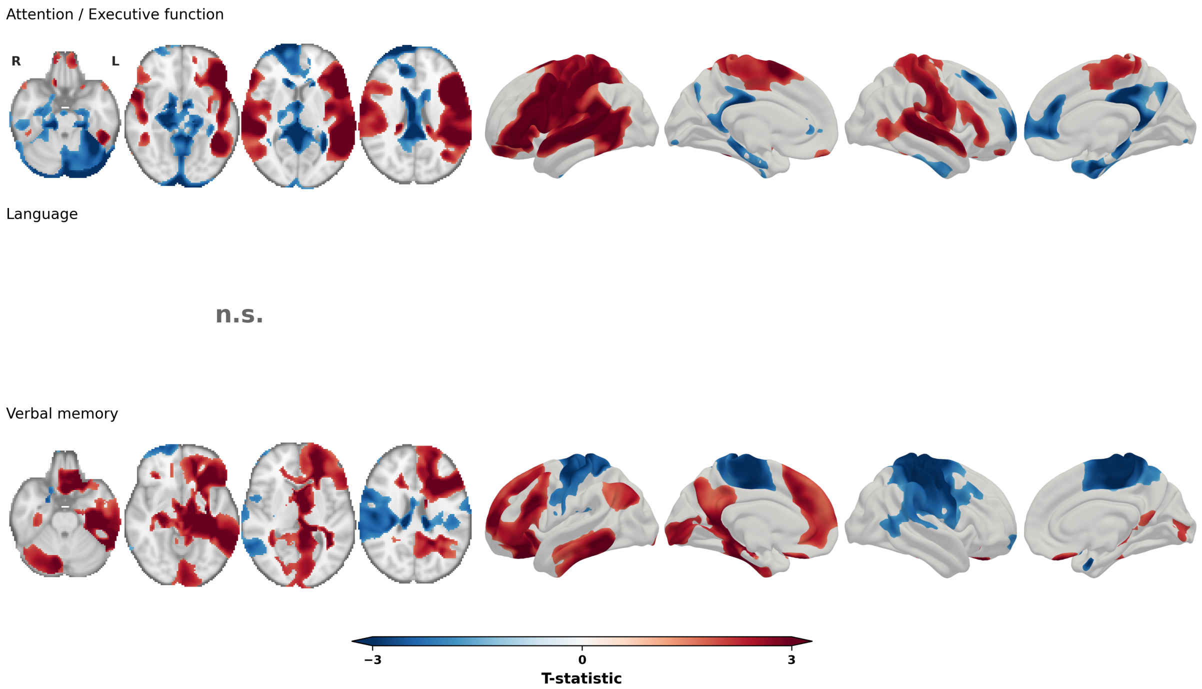


**Supplementary figure S23 | Lesion network maps adjusted for co-occurring cognitive impairments.** To test whether the similarity among the three most strongly overlapping lesion network maps reflects deficits that co-occur in the same patients, the label permutation-based analyses for attention/executive function, language, and verbal memory were repeated with the binary impairment labels of the other two domains as additional covariates (otherwise identical design and inference), restricted to patients with complete data on all three domains. Maps are thresholded at $p_{FDR} < 0.05$ (TFCE); the colour scale is capped at the 97.5th percentile of in-brain |T|. Each map retained its own topography after adjustment (voxelwise correlation with the original unthresholded lesion network map: r = 0.93 attention/executive, r = 0.81 language, r = 0.87 verbal memory), while the similarity among the three maps decreased markedly (attention/executive ~ language: r = 0.94 to 0.38, attention/executive ~ verbal memory: r = 0.57 to −0.26; language ~ verbal memory: r = 0.75 to 0.41). The adjusted language map was topographically preserved but contained no significant voxels after FDR correction. n.s. = no significant voxels.
